# Large language models identify causal genes in complex trait GWAS

**DOI:** 10.1101/2024.05.30.24308179

**Authors:** Suyash S. Shringarpure, Wei Wang, Sotiris Karagounis, Xin Wang, Anna C. Reisetter, Adam Auton, Aly A. Khan

## Abstract

Identifying causal genes at genome-wide association study (GWAS) loci remains a major challenge. Literature evidence for disease-gene co-occurrence, whether through automated approaches or human expert annotation, is one way of nominating causal genes at GWAS loci. However, current automated approaches are limited in accuracy and generalizability, and expert annotation is not scalable to hundreds of thousands of significant findings. Here, we demonstrate that large language models (LLMs) can accurately prioritize likely causal genes at GWAS loci. We rigorously evaluated several widely available general-purpose LLMs using a benchmark of high-confidence causal gene annotations, including a novel set of 26 previously unpublished GWAS. Our results show that LLMs outperform current state-of-the-art methods and substantially augment their performance. These findings establish LLMs as a powerful, efficient, and scalable approach to causal gene discovery.

## Introduction

Genome-wide association studies (GWAS) have identified numerous genomic regions associated with complex traits, enhancing our understanding of trait biology. However, pinpointing the exact causal genes within these regions remains a major hurdle. Approaches to causal gene identification from GWAS loci utilize a broad range of information including functional annotation, colocalization with quantitative trait loci (QTL) datasets, biological insights, and literature evidence. Literature mining for the co-occurrence of a (disease, gene) pair in a publication can provide evidence for the causal role of the gene in the disease and may recapitulate the knowledge that an expert biologist or clinician might use to identify the causal gene at a GWAS locus. However, current literature mining approaches^1,2^ for causal gene prioritization have been evaluated in limited settings or through indirect tasks such as drug/gene entity recognition and normalization, and their generalizability to diverse phenotypes and datasets remains unclear.

Large language models (LLMs) are deep learning models trained on large text corpora, initially to predict masked/next words from a sentence, and then subsequently trained for a large number of tasks including text generation, summarization, and question-answering. Recent studies have demonstrated their capability to perform biomedical tasks^3^, including summarizing gene function^4^, medical question answering^5^, cell-type annotation^6^, and identifying causal genetic factors from murine experimental data^7^. We hypothesize that large language models like GPT-4^8^ and Claude 3.5^9^, with extensive training on scientific publications and genetic information, provide a systematic and scalable approach to identify likely causal genes at GWAS loci. This approach has the potential to overcome the limitations of manual expert annotation.

We systematically evaluated several general-purpose LLMs for causal gene identification, comparing their performance to state-of-the-art methods (Supplementary Figure 1). Due to the difficulty in verifying LLM training sources and generalizability, we assembled three distinct evaluation sets, each with ground-truth annotations based on different criteria. The first set comprises recently published GWAS loci (after April 2023) (the “GWAS Catalog”). The second set contains loci from a benchmark not publicly available online (“Weeks et al.”). Finally, we created a new dataset of genome-wide significant loci from 26 previously unpublished GWAS from a 23andMe cohort to test performance on novel loci. Our results demonstrate that LLM-based predictions surpass existing methods, including the polygenic priority score (PoPS)^10^ and the ’nearest gene’ method^11^, in precision, recall, and F-score. Furthermore, we show that LLM predictions enhance and improve existing causal gene prioritization methods when integrated.

## Results

### LLM-based approach for causal gene identification

Our study introduces a novel approach to causal gene identification, leveraging the capacity of LLMs to analyze and synthesize information from extensive scientific literature. Unlike traditional literature mining methods, which often rely on frequency-based co-occurrence, we harness the ability of LLMs to infer complex relationships between genes and phenotypes. This approach is based on the premise that if a gene plays a causal role in a phenotype, the terms representing that gene and phenotype will exhibit strong associations within diverse scientific contexts. For instance, a specific (causal gene, phenotype) pair might have been previously discussed in publications related to a GWAS, rare disease studies, or functional experiments. Alternatively, disparate studies might exist, with some focusing on the function of the causal gene and others on the mechanisms influencing the phenotype, with known links between gene function and phenotype mechanisms. Our method uses the signal from these co-occurrences to prioritize causal genes through the knowledge encoded within the LLM.

To elicit this causal gene information, we provide each LLM with a structured text prompt that acts as an expert geneticist investigating a GWAS locus. Each prompt begins with a generic description of this scenario, followed by the specific name of the GWAS phenotype and a list of all genes located within a 500 kbp window of the lead variant (Figure 1; see Online Methods for further details). The LLM is tasked with outputting the predicted causal gene, assigning a confidence score (ranging from 0 to 1) reflecting its certainty, and providing a brief justification for its selection. Crucially, to ensure both reproducibility and transparency, we utilized available APIs to query specific, version-controlled LLM models, with detailed API usage and model information provided in the Online Methods.

**Figure 1:**
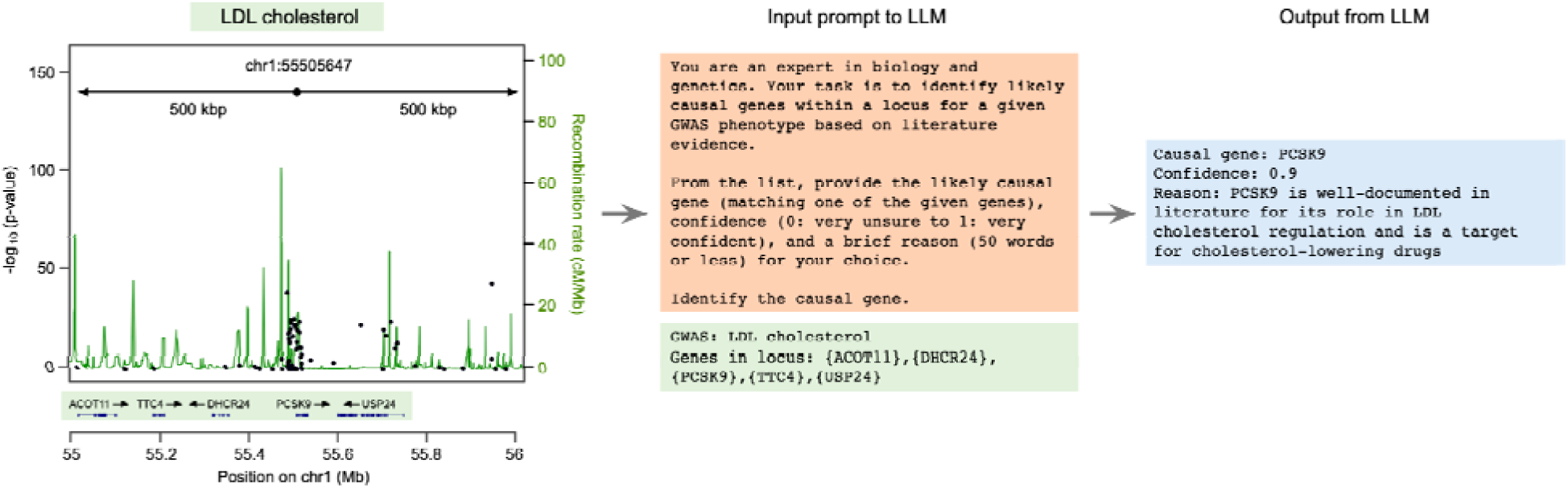
Schematic illustrating the use of LLMs for identifying causal genes at GWAS loci, with an example of an association for LDL cholesterol near the PCSK9 gene. For a GWAS locus, a 500 kbp window is extended on either side of the lead variant (indicated by the black dot), and all genes within this window are considered as candidates. The LLM is then provided with the phenotype name and the alphabetical list of genes and is instructed to predict the causal gene, its confidence, and the reason for its choice. The part of the input prompt colored in orange is the same for all loci, while the part in green is modified for each locus. For readability, output formatting instructions are excluded from the figure.

The lack of disclosure regarding the training datasets for most LLMs makes it challenging to verify whether our evaluation datasets were somehow included in their training. To mitigate this risk, we introduced a novel study design and benchmark using 641 GWAS loci recently published (after April 2023) (the “GWAS Catalog”) as well as 1,348 GWAS loci from a benchmark not publicly available online (“Weeks et al.”). We also created a new dataset of 575 unpublished GWAS loci from a 23andMe cohort to specifically test performance on novel loci (Supplementary Figure 1).

### LLMs outperform existing methods

We evaluated a range of widely available general-purpose LLMs (GPT-3.5^12^, GPT-4^8^, GPT-4o^13^, o1-mini^14^ from OpenAI; Claude 3.5 Sonnet^9^ from Anthropic; Gemini 1.5-Pro^15^ from Google, Llama 3.1-405b^16^ from Meta) for causal gene prioritization at GWAS loci. Using our prompting strategy, we observed that most recent LLMs independently demonstrated competitive performance to existing methods (Supplementary Figure 2, Supplementary Table 2). Notably, Claude 3.5 Sonnet and GPT-4o exhibited comparable performance to each other and significantly outperformed existing methods on both datasets in terms of F-score (improvements ranging from 19% to 49%, Figure 2a and 2b, Supplementary Table 2 and Supplementary Table 3). On the GWAS catalog dataset, “nearest gene” had an F-score of 0.44, while the best-performing LLM, Claude 3.5 Sonnet, achieved an F-score of 0.66. On the Weeks et al. dataset, PoPS was the best-performing non-LLM method with an F-score of 0.50, while the best-performing LLM, Claude 3.5 Sonnet, had an F-score of 0.60.

**Figure 2:**
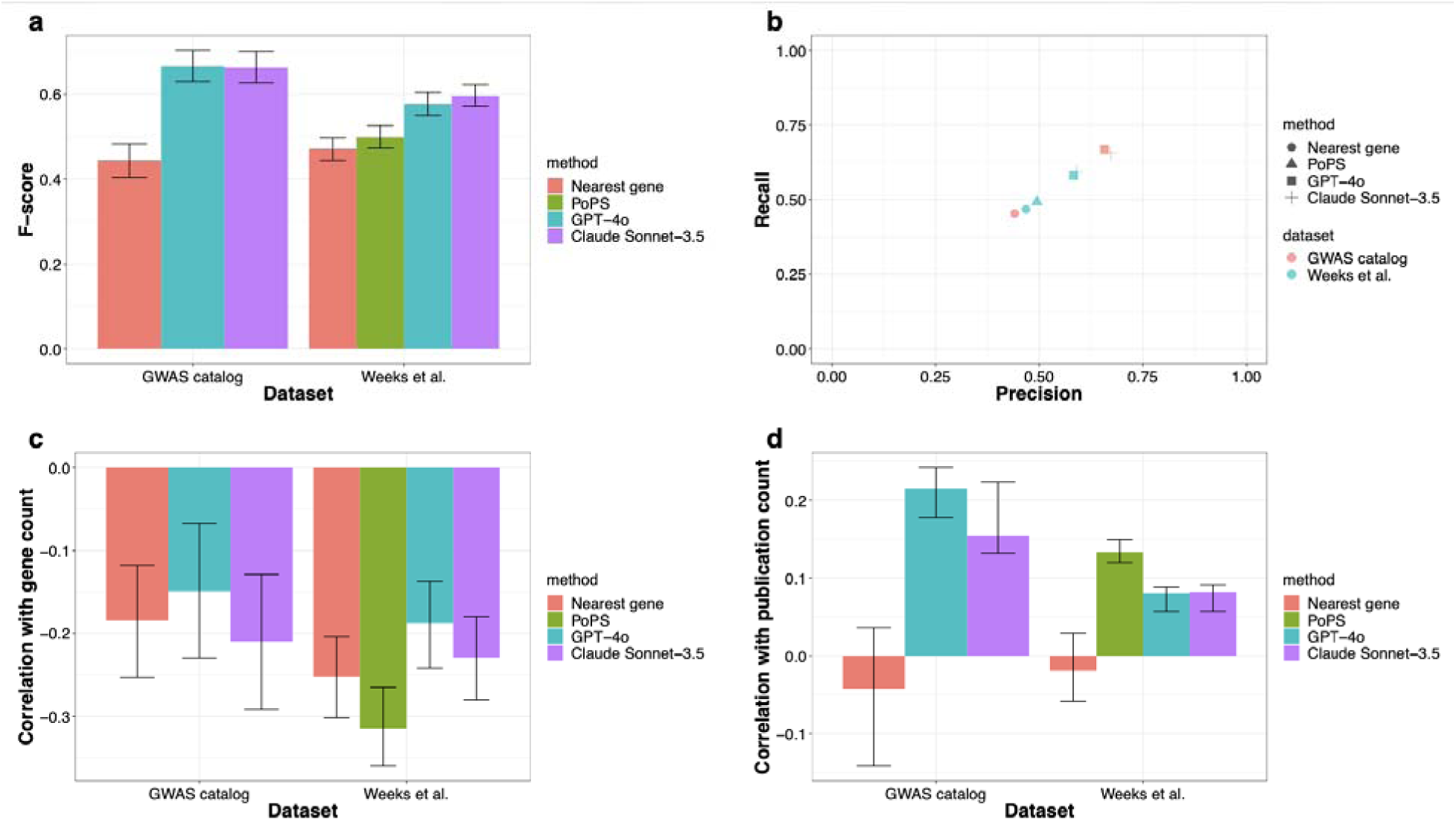
Performance of the two best-performing LLMs (Claude-3.5 Sonnet and GPT-4o) and other methods on evaluation datasets, and the factors affecting performance a. Performance of all methods on evaluation datasets as measured by F-score (a combination of precision and recall) b. Precision-recall plot showing performance of all methods on each dataset c. Impact of locus complexity on performance for all methods, measured by correlation of prediction accuracy with number of genes at locus. All methods, including LLMs, perform worse at loci with more candidate genes. d. Impact of number of publications for the causal gene on performance for all methods, measured by correlation of prediction accuracy with number of publications for causal gene. Most methods show a positive correlation of performance with how well-studied the causal gene is, except the nearest gene and L2G score methods.

To gain insights into the differential performance between methods, we investigated factors that correlated with performance. We found that the accuracy of LLM-based methods was negatively correlated with the number of genes in the locus, a relationship also observed for non-LLM methods (Figure 2c, Supplementary Table 2). Since gene-dense regions pose a challenge for causal gene prioritization methods, we stratified performance by gene density at loci, dividing each dataset into two halves using the median number of genes per locus as a threshold (Supplementary Figure 3, Supplementary Table 4). We observed a slight decrease in performance of LLM-based methods in gene-dense regions on the GWAS catalog dataset, and a more noticeable decrease on the Weeks et al. dataset (F-score decrease from ∼62% to ∼50% for both Claude 3.5 Sonnet and GPT-4o). However, non-LLM methods showed an even more pronounced drop. For instance, the PoPS F-score decreased from ∼60% on the gene-sparse subset to ∼35% on the gene-dense subset in the Weeks et al. dataset. A similar drop was observed for "nearest gene". Consequently, the advantage of LLMs over other methods is amplified in gene-dense regions, with a 38% improvement on the Weeks et al. dataset and a 67% improvement on the GWAS catalog dataset.

GWAS loci with more associated scientific literature may be expected to yield more accurate inferences of causal links to complex traits. Indeed, we also observed that LLM-based methods exhibited a positive correlation between prediction accuracy and the number of publications (as estimated from NCBI) for the causal gene. A similar correlation was observed with PoPS, which includes pathway information curated from literature evidence as a gene-level feature, but not with the "nearest gene" approach (Figure 2d, Supplementary Table 2).

### Impact of deduplication on performance

A common challenge in GWAS analysis is that a single locus may contain multiple independent signals. These signals are often close to each other, and may result in very similar or identical gene windows as input to the LLMs and other methods. To examine whether this results in an optimistic performance evaluation, we created deduplicated versions of the evaluation datasets. For each phenotype, we selected only one signal from all signals that shared the same set of genes within the 500 kbp window (Methods).

This process resulted in a reduction in dataset size from 1348 loci to 965 for the Weeks et al. dataset, and from 641 loci to 336 for the GWAS catalog dataset. We found that performance for both the LLM and non-LLM methods is reduced on both datasets, by 10-20% (Supplementary Figure 4, Supplementary Table 5). Critically, the two leading LLM methods continue to outperform the non-LLM methods even on the deduplicated datasets. In the following analyses, we use the original datasets since they more accurately result from a typical GWAS analysis followed by fine-mapping or conditional analyses.

### LLM Calibration, Reproducibility, and Reasoning

To evaluate the reliability of LLM predictions, we examined the calibration of their confidence scores. We observed that LLMs were well-calibrated at higher confidence thresholds (≥ 0.9), but tended to be overconfident at lower confidence levels (0.6–0.8). Notably, Claude 3.5 Sonnet exhibited superior calibration compared to GPT-4o (Supplementary Figure 5, Supplementary Table 6), and was therefore selected for subsequent analyses.

We next assessed the reproducibility of confidence scores by examining cases where the same prompt and predicted causal gene occurred multiple times. We found that the reported confidence scores were highly reproducible (Supplementary Table 7), with identical scores observed in 96% of instances for the Weeks et al. dataset (205 pairs) and 93% for the GWAS catalog dataset (88 pairs). This indicates that the predicted confidence is consistently derived for the same input and prediction.

To gain insights into the LLMs’ reasoning, we examined the justifications provided for their predictions. Correct predictions frequently included rationales based on gene function ("is involved in," "crucial role in," "key regulator of ") or association with the phenotype ("is associated with," "is implicated in") (Supplementary Table 8). Given the known sensitivity of LLMs to prompt structure^17^, we tested the impact of prompt variations on the models’ performance. Sensitivity analyses revealed that LLM-based methods maintained their performance even with minimal prompts that only specified the output format, task instruction, phenotype, and gene names (Supplementary Figure 6, Supplementary Table 9). This suggests that LLMs possess the knowledge required for this task, independent of the inclusion of extensive context in the prompt.

### Connection of LLM performance to representations of genes and phenotypes

To explore how genetic and phenotypic associations are internally represented, we examined the embeddings of phenotypes and genes using the “text-embedding-3-large” model^18^. LLMs represent words as points in a high-dimensional embedding space, where similarity in these representations can capture semantic relationships^19^. We hypothesized that causal genes are likely to be proximal to the phenotypes they influence in the embedding space. We tested this by calculating the cosine similarity between pre-computed embeddings of LLM-generated gene and phenotype descriptions at each GWAS locus. When using the nearest gene in embedding space from the phenotype as the predicted causal gene, we achieved approximately 58-68% of the performance of Claude 3.5 Sonnet (Supplementary Figure 7).

To illustrate this, Figure 3a presents a t-SNE projection of gene and phenotype embeddings for a locus associated with LDL cholesterol from the Weeks et al. dataset. This locus has 12 candidate genes, and the embedding of *PCSK9* is closest to the embedding of "LDL cholesterol", matching the LLM prediction. Supplementary Figure 8 shows the gene-phenotype similarity scores for this locus. Quantifying these relationships across our datasets (Figure 3b), we found that the causal gene is most similar to the phenotype in the embedding space for 40-70% of loci, depending on the evaluation set. Furthermore, the causal gene is among the top 5 most similar genes to the phenotype for 75%-93% of loci (Supplementary Figure 9).

**Figure 3:**
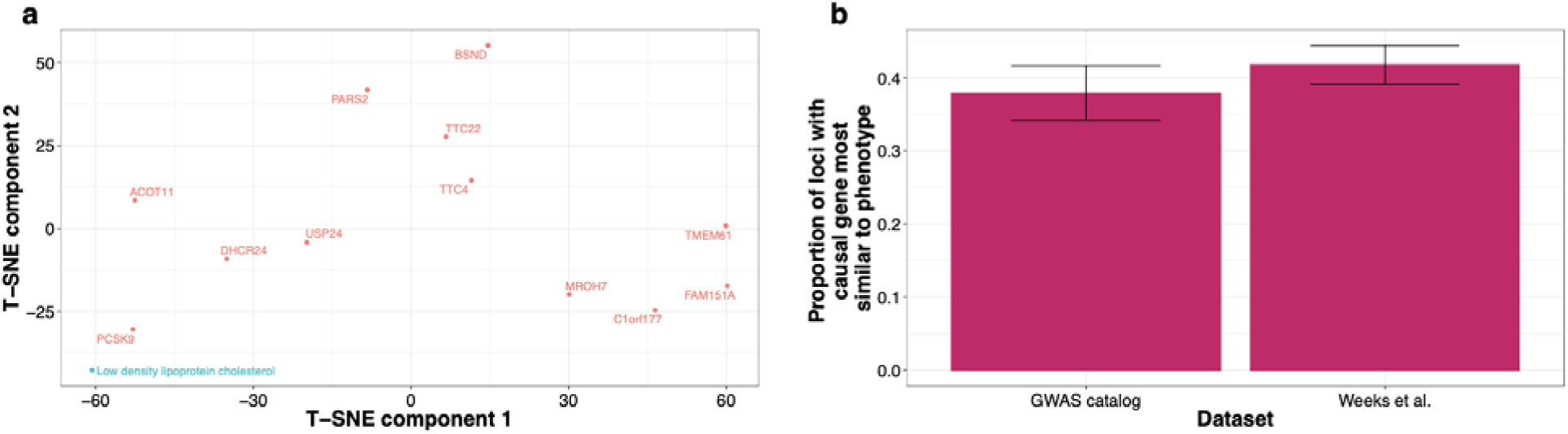
Text embeddings of genes and phenotypes partially explain performance of LLMs a) T-SNE plot to visualize text embeddings of the genes and phenotype at a locus for LDL cholesterol in the Weeks et al. data. The causal gene *PCSK9* is closest to the LDL cholesterol phenotype in the text embedding space as measured by cosine similarity. b) Proportion of examples in the evaluation datasets where the causal gene is most similar to the phenotype in the text embedding space as measured by cosine similarity.

Although embedding similarity explains a large proportion of the LLM’s performance, the context encoding performed by LLMs improves on these embeddings. For example, the GWAS catalog dataset contains 250 loci for "Multi-trait sex score", a phenotype representing sex differences and defined as a weighted sum of multiple quantitative traits. An embedding-based approach performs poorly for this phenotype (precision=0.28), whereas Claude 3.5 Sonnet achieves a precision of 0.66 (Supplementary Table 10). This indicates that, by incorporating additional context from the task description and gene information, the LLM can correctly infer that the phenotype is related to sexual dimorphism and sex-specific traits.

### Robustness of LLM Input and Reproducibility

To assess the robustness of LLM predictions to context variations, we conducted two tests with Claude 3.5 Sonnet, which exhibited superior calibration compared to GPT-4o. We evaluated performance when the causal gene was removed from the input gene list, and again when the order of genes in the input string was randomly shuffled.

Removing the causal gene from the input significantly reduced the LLM’s mean prediction confidence scores, by 0.02 for the GWAS catalog dataset and 0.07 for the Weeks et al. dataset (Supplementary Table 11). This reduction was even more pronounced for high-confidence predictions (confidence ≥ 0.9), with a 45-55% decrease in such predictions. Notably, when we specifically analyzed loci where the LLM originally made a correct prediction, the removal of the causal gene led to a 77% reduction in high-confidence predictions across both datasets.

Shuffling the order of genes in the input resulted in a match for Claude 3.5 Sonnet’s prediction at 87% of loci compared to the original, ordered input (Supplementary Table 12). Loci that received higher confidence scores from the LLM in the original dataset were more likely to produce matching predictions with shuffled inputs. Specifically, the mean confidence for matching loci was 0.1 higher for the Weeks et al. dataset and 0.06 higher for the GWAS catalog dataset compared to non-matching loci. This suggests that LLM outputs are more susceptible to input order when the LLM has lower confidence in its prediction.

LLMs generate outputs through sampling tokens from a probability distribution; the "temperature" parameter modulates this sampling variability. For reproducibility, we set temperature to 0, leading to deterministic behavior. We examined sets of loci in our evaluation datasets with identical phenotypes and gene sets. When the input was identical, Claude 3.5 Sonnet produced the same gene prediction 98% of the time for the Weeks et al. dataset (based on 209 unique input occurrences) and 99% of the time for the GWAS catalog dataset (based on 89 unique input occurrences). Reproducible outputs showed higher confidence scores than non-reproducible ones (Supplementary Table 7). These results indicate that, with temperature set to 0, LLM output is highly reproducible but not completely invariant, with a small degree of variability.

The robustness and reproducibility experiments along with the calibration results suggest an important role for LLM-predicted confidence in the interpretation of the LLM predictions - higher confidence predictions are better calibrated, more robust to input variation, and more reproducible. As we reported earlier, this predicted confidence is also highly reproducible, increasing its utility for interpreting the LLM predictions.

### LLM performance at novel loci

A critical question is whether LLMs can merely summarize existing literature about known GWAS associations, or if they possess the capacity to make accurate predictions at novel loci. To evaluate this, we used unpublished GWAS data from 23andMe, classifying loci as known or novel based on their linkage disequilibrium (LD) with index variants in the GWAS catalog (see Online Methods). Notably, our definition of novelty is stringent, as it requires that there is no prior known locus in the GWAS catalog for any phenotype that was in LD (within a pre-specified threshold) with the 23andMe hit and was unavailable to any LLM for training. Our evaluation set included 575 loci across 26 phenotypes.

We found that Claude 3.5 Sonnet’s performance on known loci was modestly better than its performance on a strictly defined set of novel loci (maximum rsquared with GWAS catalog lead variant < 0.5) (precision = 0.35 vs. 0.25), though this difference was not statistically significant (Figure 4, Supplementary Table 13). We observed a slight decrease in performance on novel loci with more lenient definitions of novelty (precision falling to 0.21 when novel loci include any with maximum rsquared < 1). These results suggest that LLMs can identify likely causal genes even at previously unreported loci.

**Figure 4.**
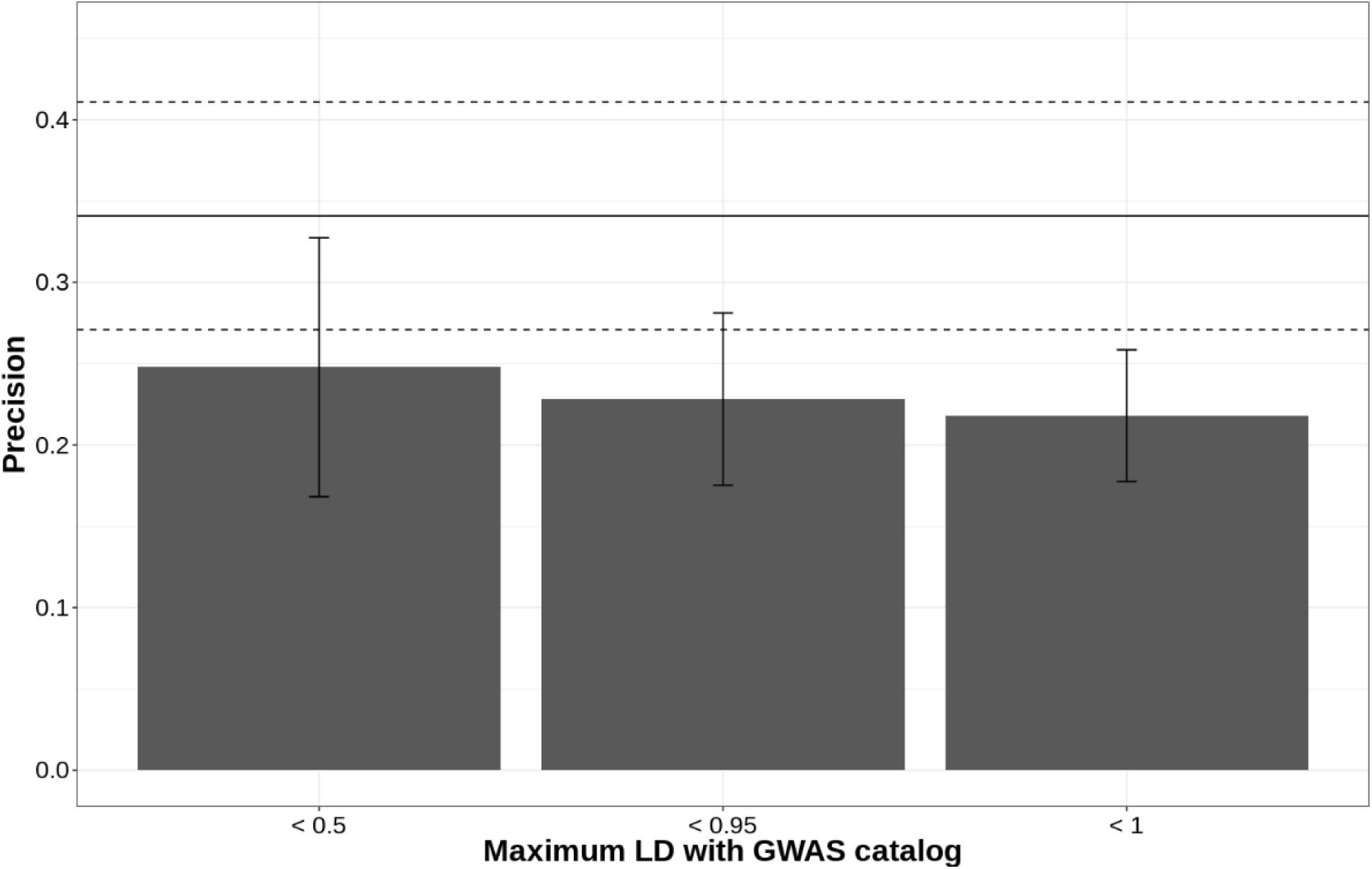
Precision of LLM predictions on evaluation set of 575 loci with linked coding variants from unpublished GWAS on a 23andMe cohort. The X-axis shows loci categorized by the maximum r-squared of the GWAS lead variant with any variant in the GWAS catalog, with increasingly lenient thresholds for the definition of novelty. The Y-axis shows precision, with 95% confidence intervals for the bars. The solid line represents the performance of LLMs at known loci, with the dashed lines indicating the 95% CI for that estimate. We see a small drop in performance for novel loci compared to known loci.

We explored whether factors previously identified as influencing LLM performance could explain this performance gap. We found that for known loci, the number of publications associated with the causal gene was significantly greater than that for novel loci (Figure 5a), which, as shown previously, correlates with LLM performance. Consequently, LLM predictions at known loci had higher average confidence scores than predictions at novel loci (Figure 5b).

**Figure 5:**
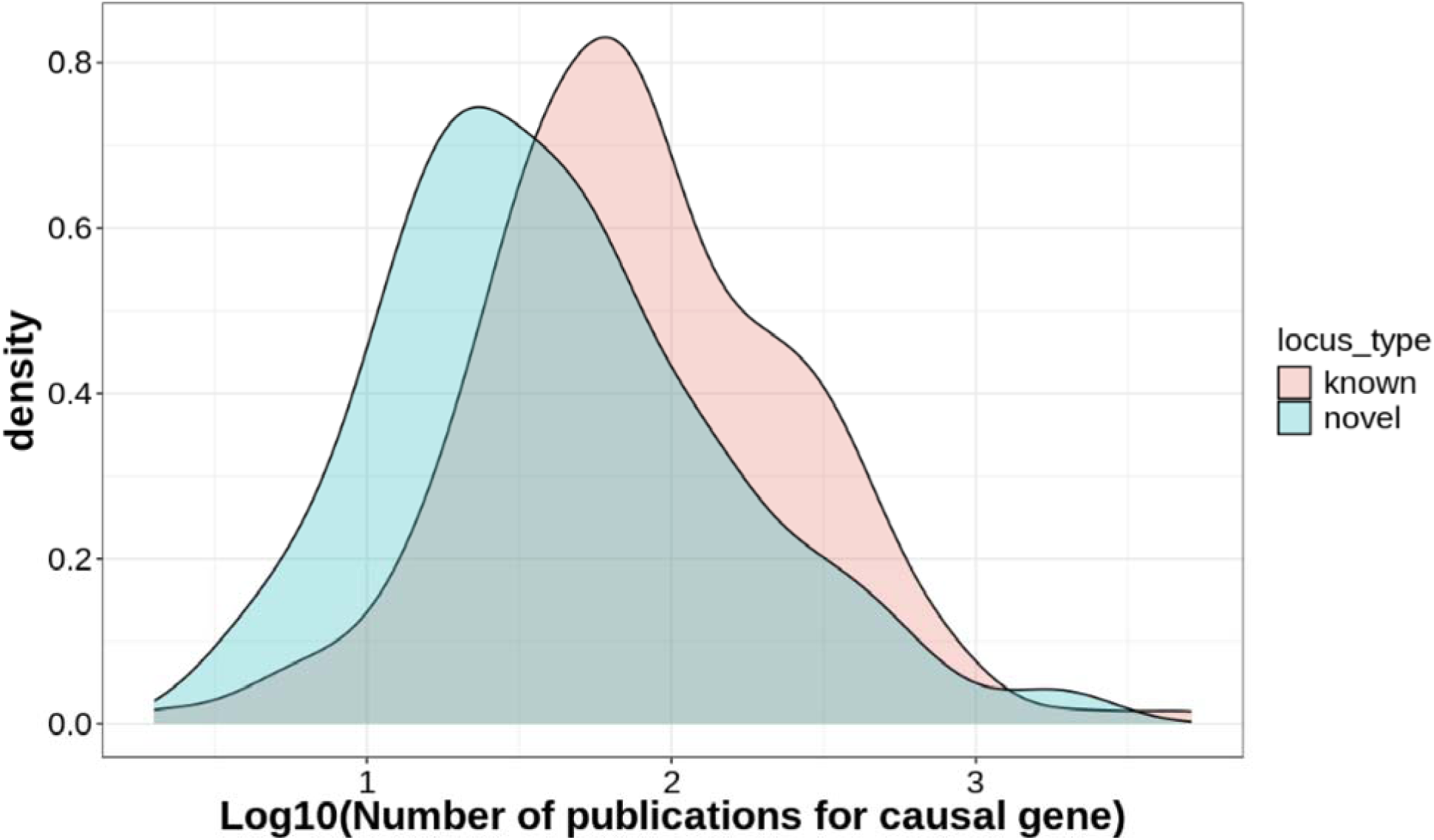
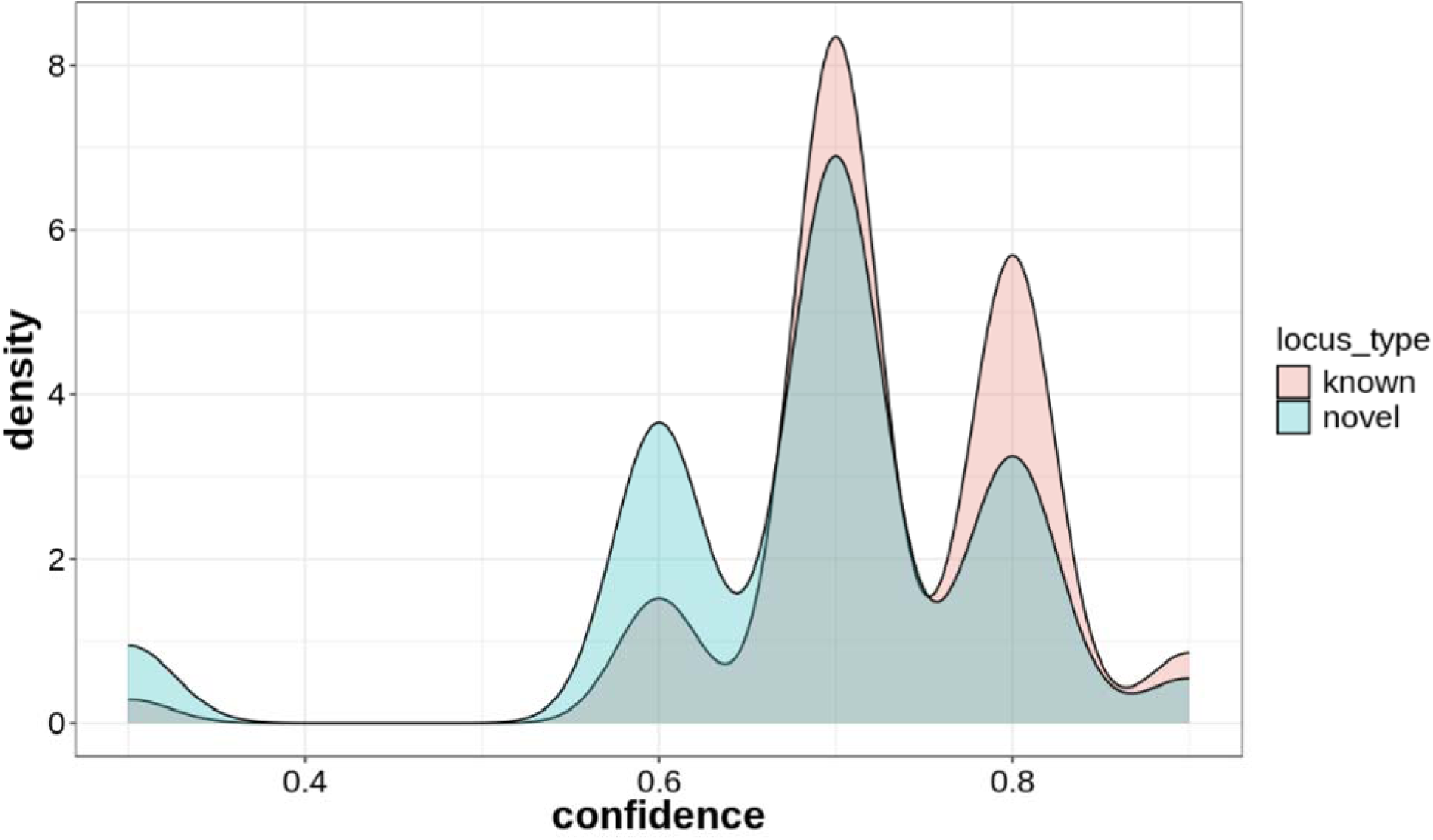
Differences between known and novel loci (novel here means the lead variant has maximum LD less than 0.5 with any hit in the GWAS catalog) (a) Number of publications for the causal genes at known vs novel loci. The number of publications for causal genes at known loci is larger than for causal genes at novel loci.(b) Confidence of LLM predictions at known vs novel loci. The LLM makes more high-confidence predictions for known loci than novel loci.

These findings suggest that the observed performance gap between known and novel loci may be correlated with the LLM’s predicted confidence. Therefore, we performed a similar analysis on a larger dataset, encompassing more phenotypes and loci. With a larger sample size, we observed similar aggregate results (Supplementary Figure 10a). However, when we stratified the results by the LLM-predicted confidence, we found that the performance gap between known and novel loci was significantly reduced (Supplementary Figure 10b).

### Failure modes of LLMs

We identified a few of notable failure modes by examining phenotypes where LLMs exhibited low precision and instances where newer LLMs avoided such failures. For example, GPT-4- 0613 achieved a precision of only 0.08 for the "Total protein" phenotype. The LLM’s justifications suggest an incomplete understanding of the short phenotype description, interpreting it broadly in terms of general protein levels rather than specifically about protein levels in blood. This indicates that more specific phenotype descriptions could enhance LLM performance. However, newer LLMs such as Claude 3.5 Sonnet (and GPT-4o) did not demonstrate similar issues with this phenotype, achieving a precision of 0.52.

In another instance, Claude 3.5 Sonnet had a precision of 0.0 for the phenotype "Neonatal circulating Complement Component 4 (C4) protein concentration." For this phenotype, all GWAS loci in the evaluation dataset included the *C4A* gene within the 500 kbp window of the lead variant, despite these loci having different causal genes based on coding variant signals. Because the *C4A* gene is a major component of the complement system, the LLM-based approach consistently predicted *C4A* as the causal gene. This highlights the potential for combining LLMs with functional annotation data to improve causal gene prioritization. Supplementary Table 10 shows the precision and recall for all methods stratified by phenotype.

Finally, we observed a small number of hallucinations, where the LLMs reported a causal gene not included in the set of provided genes at a locus (fewer than 4% of all loci for Claude 3.5 Sonnet, fewer than 1% for GPT-4o, Supplementary Table 14). We also tested “chain-of-thought” prompting^16^ to see if it might reduce the low hallucination rate, but found no improvement in performance (Supplementary Figure 11).

### Combining LLMs and other gene prioritization methods

We sought to investigate whether LLMs provide orthogonal information to existing gene prioritization methods and whether combining them would enhance causal gene prediction. Previous work has shown that combining multiple gene prioritization methods improves performance^10,11,20^. We hypothesized this would also apply to LLM-based approaches. We began by examining the concordance of predictions across different methods. We found that LLM-based methods showed the highest agreement with other LLM-based methods, but only moderate agreement with the polygenic priority score (PoPS) and the ’nearest gene’ methods (Supplementary Figure 12, Supplementary Table 15). These findings suggest that LLMs and existing methods capture distinct aspects of the data, implying a potential for improved performance through combined approaches.

To explore this potential, we combined LLM-based approaches with other gene prioritization methods. First, we considered a simple consensus-based prediction approach, using only concordant predictions by LLM and other methods while discarding discordant predictions. We found that this consensus approach improves precision but at the cost of decreasing recall due to the removal of discordant predictions (Figure 6). The precision of consensus prediction using LLMs and one other non-LLM method ranged from 0.86 on the Weeks et al. dataset (nearest gene + Claude 3.5 Sonnet) to 0.89 on the GWAS catalog data (nearest gene + either LLM), with corresponding recall values of 0.33 and 0.36, respectively.

**Figure 6:**
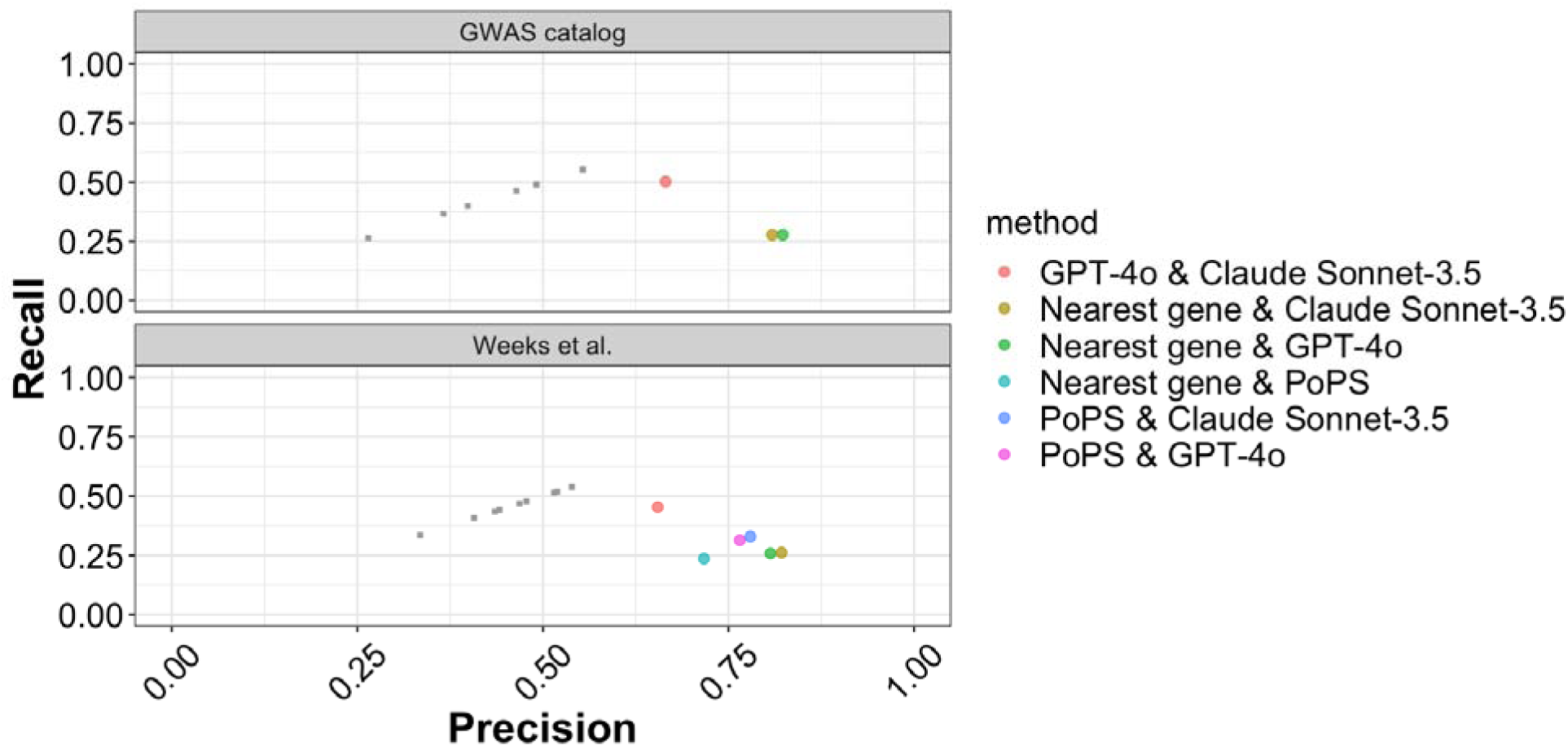
Performance of consensus prediction methods across all datasets, depicting precision and recall. Gray squares represent the performance of individual methods. Consensus prediction methods consistently exhibit higher precision but lower recall compared to individual methods.

The observed trade-off between precision and recall with a consensus-based approach highlighted the need for a framework that could prioritize predictions in cases of disagreement. To address this, we developed an ensemble decision tree classifier (Methods) trained to select the most likely accurate prediction for a given locus. This classifier leverages both the agreement between methods and the confidence score provided by the LLM. In this way, the classifier learns highly interpretable rules to select predictions from different methods to predict a causal gene.

To train our classifier, we used agreement between methods and the LLM-predicted confidence as features. Figure 7a demonstrates the performance of an ensemble classifier created using predictions from the “nearest gene” method and Claude 3.5 Sonnet. This predictor significantly outperformed the ’nearest gene’ method alone across all datasets, with F-score improvements ranging from 29% (from 0.47 to 0.61) on the Weeks et al. dataset to 48% (from 0.45 to 0.67) on the GWAS catalog dataset (Supplementary Table 16). Similarly, combining LLM predictions with those from the PoPS method resulted in notable improvements (Figure 7b), with a 17% increase in F-score (from 0.50 to 0.59) over PoPS alone on the Weeks et al. dataset (Supplementary Table 17).

**Figure 7:**
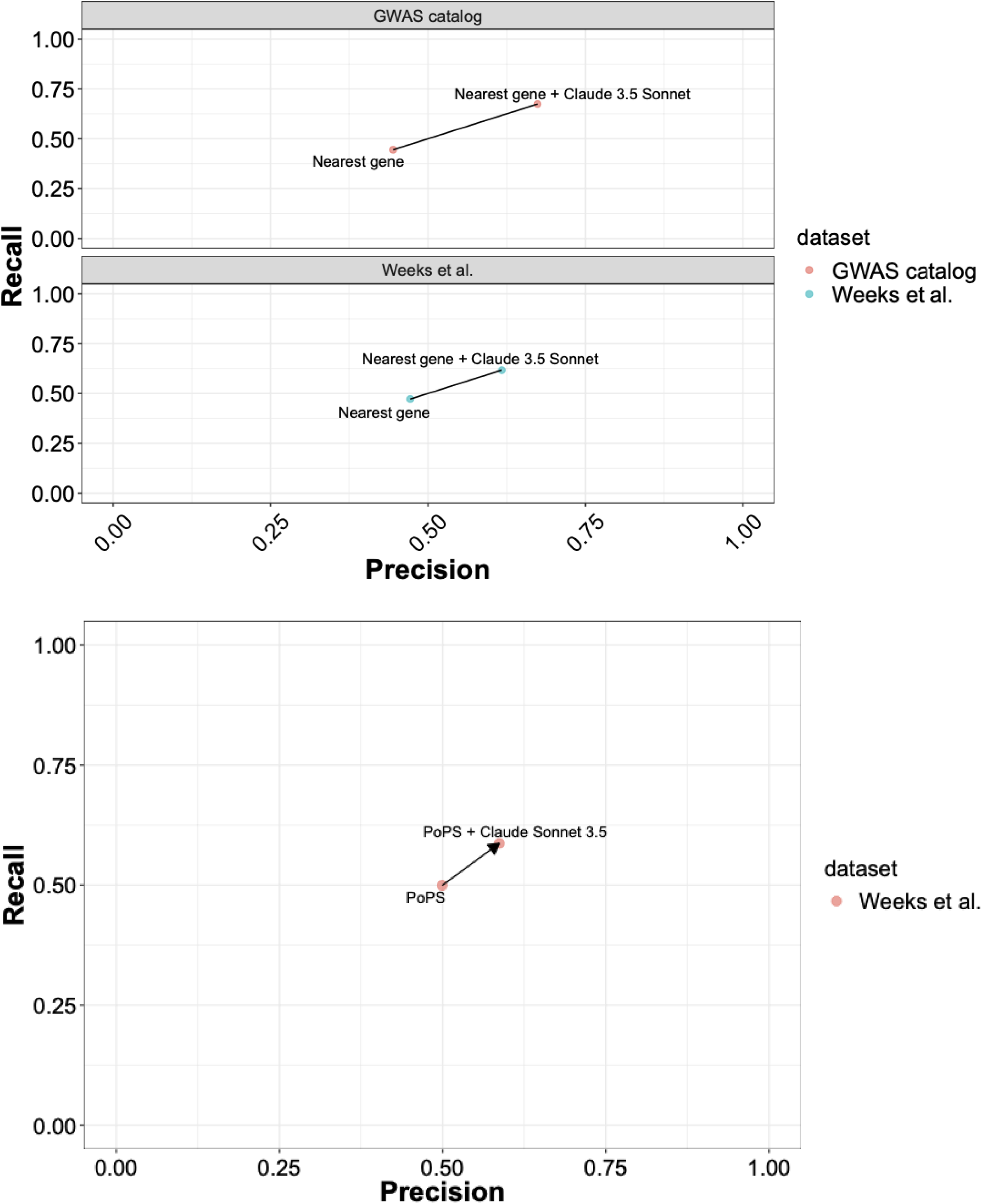
Performance comparison of ensemble framework to individual methods with respect to precision and recall. Comparison of ensemble predictions from Claude Sonnet-3.5 and Nearest gene against predictions from Nearest gene alone for the GWAS catalog and Weeks et al. datasets

The ensemble framework is highly interpretable and can be described using simple, learned rules (Methods). When both methods agree, the decision tree selects the consensus prediction. However, when methods disagree, the decision tree selects the LLM prediction if the LLM confidence score is high; otherwise, it defaults to the prediction from the other method (Supplementary Table 18). These results demonstrate that integrating the unique information from LLMs with traditional methods offers a powerful approach to enhance causal gene prediction.

### Analysis costs for LLM usage

The LLMs employed in our analyses are predominantly closed-source and only accessible through paid APIs. Usage costs for API-accessed models are based on the size of the input query and the size of the output, measured in number of “tokens” (words or combinations of characters). Costs are also variable based on the model. We found that the average per-locus cost for Claude 3.5 Sonnet was $0.0022 USD for the GWAS catalog dataset, and $0.0021 USD for the Weeks et al. dataset. Other methods had similar costs, with the exception of the OpenAI o1-mini model, where costs were higher due to hidden reasoning tokens. As an example, the complete annotation for GWAS with 300 loci can be computed for less than $1 USD.

## Discussion

Our study demonstrates for the first time that LLMs significantly enhance causal gene identification and GWAS interpretation by systematically integrating literature evidence. Our approach, requiring only the lead variant’s genomic location, offers broad applicability to any GWAS locus, circumventing the limitations of incomplete summary statistics prevalent in many studies. Critically, our method outperforms traditional approaches when applied to an evaluation dataset created from the widely used GWAS catalog resource, where less than half of the studies included in the dataset provided summary statistics. Unlike existing literature-based methods^1,2^ that rely on phenotype-to-ontology mappings, our approach mitigates difficulties arising from ambiguous or non-standard phenotypes. A key feature of our method is the use of a per-locus confidence score, a crucial metric for result interpretation, reproducibility, and calibration that is often lacking in state-of-the-art methods.

Using unpublished GWAS loci from a 23andMe cohort, we further show that LLMs identify likely causal genes at novel loci, albeit with potentially reduced performance compared to well-studied loci. This suggests that LLMs do not simply retrieve known gene-phenotype associations, but can infer new relationships. Moreover, an ensemble approach combining LLMs with established methods substantially improves causal gene identification.

While we have demonstrated the potential of LLMs for causal gene identification, it is important to consider the limitations and challenges associated with their use in our study. The lack of transparency in LLM training datasets raises concerns about potential training data contamination. As an example, we found that LLM performance was inflated on the Opentargets and Pharmaprojects datasets, which have been previously used as gold-standard causal gene datasets, compared to the Weeks et al. and GWAS catalog dataset (Supplementary Figure 13). One reason for this could be the availability of these datasets on the internet. Therefore, we excluded these datasets from our evaluation.

To further mitigate the risk of LLMs being trained on evaluation datasets, we introduced a study design using data curated after known LLM training periods as well as a benchmark dataset not available on the internet. Additionally, we conducted checks by applying a heuristic method^21^ to determine if the LLMs had ingested any of the evaluation datasets as structured tables, and found no evidence of contamination. While LLMs return plausible reasons along with their predictions, it is challenging to pinpoint the exact information the LLM used to make each prediction. Including citations in the response could reduce this problem, but prompting current LLMs to include citations in their response is prone to hallucination, and not yet useful. This is a current shortcoming, but it also points to an area of active research in the field.

Given the rapid pace of development in LLMs, several practical considerations remain for their application to causal gene prioritization. Real-world implementation necessitates careful attention to data requirements, computational costs, and the inherent trade-offs between performance and transparency. Our study used a 500 kbp genomic window for causal gene prediction, a standard distance used in causal gene prioritization methods^10,11,20^ and modifiable by users. Furthermore, we anticipate that advancements in LLM reasoning capabilities, including the use of Retrieval-Augmented Generation (RAG) and agent-based systems, will improve interpretability. Although internet access is becoming increasingly common in LLMs for additional context retrieval, we did not explore these features due to concerns about reproducibility and interpretability. Finally, while we estimate a cost of approximately $1 USD for a GWAS with 450 significant loci, pricing and other cost factors may change, especially as more complex reasoning-based LLM services and other technologies emerge.

With LLMs continuously updating with larger training data, these widely available general-purpose LLMs offer an efficient and scalable way to harness expanding scientific literature and knowledge for improved causal gene identification. Ultimately, combining LLM-based approaches with functional or other multimodal data holds the promise of creating even more accurate and robust methods for identifying causal genes, thereby advancing our understanding of genetic contributions to complex traits.

## Data Availability

We will share all processed datasets used in our analysis, as well as the prediction results from all methods on all datasets, intermediate outputs like gene and phenotype embeddings using Zenodo (doi: 10.5281/zenodo.11391053).

## Methods

### Evaluation datasets

We used two datasets for evaluation of all methods (1) an evaluation set created by Weeks et al. based on proximity to fine-mapped coding variant associations, and (2) an evaluation set we created using associations added to the GWAS catalog from manuscripts published after April 2023.

In initial experiments, we used two additional datasets from OpenTargets and Pharmaprojects that have been previously used in causal gene prioritization work. However, we found that LLMs had inflated performance on them compared to the other two, likely because the OpenTargets and Pharmaprojects data are available on the internet and may have been used for LLM training.

The Weeks et al. evaluation set contains non-coding credible sets that are within 500 kbp of a high-confidence (posterior inclusion probability PIP > 0.5) fine-mapped coding association in the same GWAS in UK Biobank^10^. In this dataset, the authors considered pairs of credible sets that were within the same GWAS. If one of the credible sets was composed entirely of non-coding variants and the other contained a high-confidence coding variant within 500 kbp of the first credible set, then the authors considered the lead variant of the non-coding credible set as the GWAS hit of interest, and the causal gene was the gene to which the coding variant belonged. We obtained this dataset by emailing the authors. Since this dataset is not available directly on the internet, it is the least likely to have been directly used for training the LLMs evaluated in our study (though the underlying GWAS summary statistics and publication could be part of the LLM training data). We used the gene symbol annotations provided by the authors. This resulted in a dataset with 1348 causal gene - phenotype pairs.

The GWAS catalog dataset contains non-coding lead variants that are within 500 kbp of a coding lead variant in the same GWAS study. We downloaded the GWAS catalog lead variant file version “gwas_catalog_v1.0.2-associations_e111_r2024-03-11.tsv” (download date: 03/19/2024). To minimize the possibility of these results being included in LLM training datasets, we subset the data to only include associations added to the GWAS catalog for manuscripts published after April 30, 2023. To assign causal genes at loci, we considered loci with non-coding lead variants. If this lead variant was within 500 kbp of a coding lead variant in the same study, we include the non-coding locus in our dataset, with the gene that the coding lead variant belongs to as the causal gene. This resulted in a dataset with 641 causal gene - phenotype pairs.

We tested whether any of our evaluation datasets were directly included in the LLM training datasets, and did not find evidence for that. Details of the testing procedure are included in Supplementary Note (Section: Contamination checks), and quantitative results are included in Supplementary Table 19.

### Generating LLM input from datasets

For input to the LLMs, we converted each (phenotype, lead variant, causal gene) triplet corresponding to a GWAS locus to a (phenotype, list of genes in locus) pair.

For OpenTargets, Pharmaprojects, and the GWAS catalog datasets, we identified all genes within 500 kbp of the lead variant (using GENCODE release 43) based on the smallest distance between the gene body and the lead variant. For the Weeks et al. dataset, the dataset provided by the authors included all genes within 500 kbp of the lead variant, so we used that list of genes without any additional processing.

To avoid leaking information about the lead variant location or causal gene position through the ordering of gene symbols by physical position, we sorted all gene symbols lexicographically before including them in the prompt to the LLMs.

As an additional sanity check to verify that the LLM relies on phenotype information and not just on gene information (such as location) for predictions, we randomly sampled phenotypes at the evaluation loci and provided those as input to Claude 3.5 Sonnet. As expected, we found that LLM performance is considerably degraded from an average precision of 63% across all datasets to an average precision of 34%, along with an increase in the number of obvious hallucinations from 22 to 45 (Supplementary Table 20).

### LLM prompts for identifying causal genes

To query LLMs to identify the causal genes for a (phenotype, list of genes in locus) pair, we used a prompt describing the task for the LLM along with the phenotype and gene list. The basic prompt we used had two components, a system prompt (for general behavior), and a user prompt (pair-specific). For an example pair (“Morning person”,[A,B,C,D]), the prompts are described below.

System prompt:

You are an expert in biology and genetics.

Your task is to identify likely causal genes within a locus for a given GWAS phenotype based on literature evidence.

From the list, provide the likely causal gene (matching one of the given genes), confidence (0: very unsure to 1: very confident), and a brief reason (50 words or less) for your choice.

Return your response in JSON format, excluding the GWAS phenotype name and gene list in the locus. JSON keys should be ’causal_gene’,’confidence’,’reason’.

Your response must start with ’{’ and end with ’}’.

User prompt:

Identify the causal gene.

GWAS phenotype: {Morning person} Genes in locus: {A},{B},{C},{D}

### LLMs evaluated

For our experiments, we evaluated LLMs from OpenAI (GPT-3.5, GPT-4, GPT-4o, o1-mini), Anthropic (Claude 3.5 Sonnet), Google (Gemini 1.5 Pro), Meta (Llama 3.1-405b). For GPT-3.5, we used the model version “gpt-3.5-turbo-0125”, which has been trained on data up to September 2021. For GPT-4, we tested the model version “gpt-4-0613”, which has been trained on data up to September 2021. For GPT-4o, we used model version “gpt-4o-2024-08-06”. We tested o1-mini in the o1 series for latency and cost reasons, and because the model was optimized for scientific reasoning. For the Claude series of models, we used V1 of the Claude 3.5 Sonnet series, and found little difference when testing V2 (results not shown). We used the largest Meta Llama 3.1 model available, expecting it to have the best performance in that series of models.

To make outputs reproducible, we queried all LLMs with temperature set to 0. We queried the OpenAI and Google models through their respective APIs using python scripts (Code availability). To access models by Anthropic and Meta, we used AWS Bedrock with Langchain^22^.

### LLM Hallucinations

We consider an LLM prediction to be an obvious hallucination if the predicted gene was not in the list of genes at the locus provided to the LLM. These are easily detectable with a string match. For a conservative evaluation, we penalize such predictions as incorrect in our analyses. In practical applications, users can easily detect these and set them to NA to improve predictions.

### Comparison to other methods

We compared our results to state-of-the-art methods for both datasets. First, we evaluated the “nearest gene” method for all datasets. We added prediction based on polygenic priority score (PoPS) to the Weeks et al. evaluation dataset. PoPS was previously evaluated to have good performance on the Weeks et al. dataset.

To obtain the nearest gene prediction, we computed the distance between protein-coding genes in each locus and the lead variant (using GENCODE release 43 and reference genome hg38 for the GWAS catalog datasets), and defined the nearest gene as the gene with the least distance from the lead variant based on gene body. For the Weeks et al. data, we directly used the nearest gene predictions provided by the authors (downloaded from https://www.finucanelab.org/data, file: https://www.dropbox.com/sh/o6t5jprvxb8b500/AACqCux_jJbF9F56ozhzzkpia/results/UKB_AllMethods_GenePrioritization.txt.gz?dl=0). In the Weeks et al. data, we found that “nearest gene” nominated multiple genes at about 4% of loci (51 of 1348) due to the lead variant position being within multiple gene bodies. To simplify evaluation, we randomly chose a single gene out of all nominated genes as the prediction at such loci. We found that this had a minor impact on precision and recall compared to that reported in the original publication (precision = 0.47 vs previously reported 0.46, recall = 0.47 vs previously reported 0.48).

PoPS is a similarity-based gene prioritization method that leverages gene-level summary statistics and incorporates data about genes from a variety of sources to produce a phenotype-gene level prioritization score. We obtained PoPS scores for the 1348 evaluation loci by emailing the authors of the original publication^10^. The predicted causal gene based on the PoPS score was defined as the gene with the highest PoPS score in the locus.

### Evaluation of predictions

For evaluation of predictions, we computed precision (proportion of predicted causal genes that were annotated as causal in the dataset), recall (proportion of annotated causal genes identified among predictions), and F-score (harmonic mean of precision and recall). All methods (nearest gene, PoPS, LLM-based approaches) made a single causal gene prediction for each example. To compute these metrics, the predictions from each method were converted to 0/1, with 1 assigned if the method made a prediction and it matched the annotated causal gene, 0 assigned if the method made a prediction but it did not match the annotated causal gene.

Precision = (Number of predictions scored 1) / (Number of non-NA predictions)

Recall = (Number of predictions scored 1) / (Number of examples in dataset)

F-score = 2 * Recall * Precision / (Recall + Precision)

For each metric, we computed 95% confidence intervals using the bootstrap with 1,000 samples.

To compare the performance of LLMs to the best non-LLM approach, we used a Wilcoxon signed rank test with continuity correction, and we report p-values for the alternative hypothesis that the LLM performance is higher than the highest non-LLM approach performance. We also report a p-value from a McNemar test for the same comparison with the alternative hypothesis of a difference between performance for the two methods. We used a similar approach when comparing Claude 3.5 Sonnet to GPT-4o.

### Factors affecting prediction accuracy

To assess the impact of locus complexity, we used the number of genes in the locus as a measure of its complexity, and computed the Pearson correlation between the number of genes in the locus and whether the prediction was correct. To stratify the performance on the gene-sparse and gene-dense subsets, we used a dataset-specific threshold to divide each dataset into two. Gene-sparse loci were defined as the loci where the number of genes in the locus was equal to or less than the median number of genes per locus across loci in the dataset. Gene-dense loci were defined as the remaining loci, where the number of genes in the locus was larger than the dataset median.

To assess the impact of publication count, we used publication counts for causal genes to evaluate correlations with prediction performance. We downloaded the “gene2pubmed” file from https://ftp.ncbi.nlm.nih.gov/gene/DATA/ (download date: January 26, 2024). We subset the file to only include human genes (tax_id = 9606), and count the number of publications per gene. Since many examples might share the same causal gene, for each causal gene, we computed the proportion of loci sharing the same causal gene predicted correctly. To account for the heavy-tailed distribution of publication count by gene, we computed the Spearman correlation between the number of publications for the causal gene and the proportion of correct predictions at loci with the specified causal gene.

We used the bootstrap with 1,000 samples to compute 95% confidence intervals for both correlations. We used the same resampled dataset for confidence interval calculations for precision, recall, F-score and both correlations.

### Calibration analysis and confidence reproducibility

To assess calibration, we used the confidence scores provided by the LLM. We computed precision for all predictions at a given confidence score, for any confidence score with at least 5 predictions. Supplementary Table 6 shows all unique confidence scores predicted by the LLMs, with their counts and precision estimates. We computed a standard error for each precision by assuming it to be the mean of a binomial distribution with size given by the number of predictions with that score. 95% confidence intervals were calculated as precision +/- 1.96*se.

To evaluate the reproducibility of the confidence metric, we considered loci in each dataset which shared the same (prompt, predicted causal gene) pair and appeared multiple times. For each such locus, we calculated the proportion of times the LLM predicted exactly the same confidence value.

### Summarizing reasons provided by the LLM for correct predictions

Across all datasets, the LLMs make hundreds of correct predictions. To summarize these for easy inspection, we concatenated all reason strings provided by Claude 3.5 Sonnet for predictions evaluated to be correct. We then identified trigrams in the concatenated string using the ngram package in R, and sorted them in decreasing order of frequency of occurrence to identify the most common ones. We report the top 200 most frequent trigrams in Supplementary Table 8.

### Sensitivity analysis for prompt structure

To examine how the prompt affects the prediction accuracy of the LLMs, we experimented with replacing the prompt by a minimal alternative containing only the output format, task description, and the locus information. We conducted these experiments with Claude 3.5 Sonnet. The detailed prompt is described below, using the same example locus as earlier.

System prompt:

From the list, provide the likely causal gene (matching one of the given genes), confidence (0: very unsure to 1: very confident), and a brief reason (50 words or less) for your choice.

Return your response in JSON format, excluding the GWAS phenotype name and gene list in the locus. JSON keys should be ’causal_gene’,’confidence’,’reason’.

Your response must start with ’{’ and end with ’}’.

User prompt:

Identify the causal gene.

GWAS phenotype: {Morning person}

Genes in locus: {A},{B},{C},{D}

### Embeddings for genes and phenotypes

We generated descriptions for genes and phenotypes using GPT-3.5 (model version “gpt-3.5-turbo-0125”) by using the prompt below.

"You are an expert in biology and genetics.

Your task is to provide biologically relevant information about the query below in 300 words or less.

Query: {text}."

The “text” value was generated by adding the entity type (gene/phenotype) to the beginning of the entity. So the value of text for the *BRCA1* gene would be “gene *BRCA1*”, and that for the breast cancer phenotype would be “phenotype breast cancer”.

We provided the generated descriptions as input text to get embeddings using the OpenAI text embedding model “text-embedding-3-large”. This produced embeddings with 3,072 dimensions.

### Embedding-based causal gene prediction

Using the computed embeddings, we provided the embedding-based causal gene prediction. We calculated the dot product (identical to cosine similarity since the embeddings are standardized) between the phenotype embedding and the gene embedding, and we defined the predicted causal gene as the gene with the largest dot product of phenotype and gene embeddings among all the genes in each loci.

To create the t-SNE plot for the *PCSK9* locus, we ran t-SNE on the 13 data points (1 phenotype + 12 genes in the locus) using the Rtsne package with a perplexity of 4.

### LLM robustness and reproducibility

To test the robustness of LLMs to input which does not include the causal gene, we removed the causal gene from the input list of genes included in the LLM prompt (the “null dataset”). We compared the confidence between the original dataset and null dataset to see if the LLM predictions were affected. To test the effect of gene order, we randomly shuffled the input gene list in the LLM prompt, and compared the predictions for the two datasets. We use the confidence from the original prediction to examine correlation with whether the output changes.

To examine reproducibility of causal gene predictions, we considered loci in each dataset which had the same input gene list and appeared multiple times. For each such locus, we calculated the proportion of times the LLM predicted the same causal gene.

### LLM performance at novel loci

For this evaluation, we used unpublished GWAS based on 23andMe data to develop an evaluation set using loci where the lead variant is either a coding variant or in strong LD (rsquared >= 0.9) with a coding variant. We considered a locus as known if the lead variant was either directly present in the GWAS catalog or in perfect LD (rsquared = 1.0) with a variant in the GWAS catalog. Note that this is a strong definition of novelty since we do not require the GWAS catalog phenotype to be the same as the 23andMe phenotype. We considered a locus novel if it was not in strong LD with any hit in the GWAS catalog (maximum rsquared below a variable threshold). Our evaluation set included 575 loci from 26 phenotypes.

### LLM failure modes

To better understand LLM failures, we computed precision by phenotype. To implement chain-of-thought prompting, we used Claude 3.5 Sonnet along with DSPy^23^.

### Ensembling LLM predictions with non-LLM predictions

To ensemble LLM predictions with non-LLM predictions at different loci in the evaluation datasets, we framed the problem as a multi-label classification problem, to predict whether each individual method makes the right prediction at a locus. For each locus and method, we want the ensemble classifier to predict its most likely label (correct/incorrect), and the associated probability that the prediction is correct. Then, the ensemble prediction is the prediction from the method that has the highest probability of having the right prediction. We used a decision tree as our classifier.

To avoid data leakage during training the ensemble classifier, we used a nested cross-validation setup, where loci from a specific chromosome are assigned to a single outer cross-validation fold, and for predictions for these loci, we use a model that is trained by excluding this outer fold. We use standard cross-validation within each training fold to identify the best decision tree depth and number of samples to split a node.

We used the scikit-learn package^24^ to implement the ensemble predictor.

### Examining ensemble predictions

To examine ensemble predictions, we generated all possible feature combinations of the confidence feature and agreement feature. The confidence feature took values from {0.7, 0.8, 0.9, 1.0} to match observed values, and the agreement feature was a binary indicator. For the interpretation task, the decision tree was trained on the full dataset (instead of the nested cross-validation design), and we used the trained classifier on every possible feature combination.

## Supplementary Tables

Table 1 - Datasets

Table 2 - Performance for all methods

Table 3 - Pairwise test LLM vs non-LLM

Table 4 - Performance of methods in gene-dense and gene-sparse regions

Table 5 - Performance of methods on deduplicated datasets

Table 6 - Calibration of LLM predicted confidence

Table 7 - Reproducibility of predictions and confidence

Table 8 - LLM reasons

Table 9 - Ablation study

Table 10 - LLM precision by phenotype

Table 11 - Causal gene deletion result

Table 12 - Input gene order shuffling result

Table 13 - Performance on novel vs known loci

Table 14 - Hallucinations

Table 15 - Pairwise concordance of methods

Table 16 - Ensemble of nearest gene + LLM

Table 17 - Ensemble of PoPS + LLM

Table 18 - Ensemble method predictions for all feature combinations

Table 19 - Contamination checks

Table 20 - Performance on shuffled phenotypes

## Acknowledgements

The authors thank the employees and research participants of 23andMe for making this work possible. We acknowledge Elle Weeks, Hilary Finucane, and Jacob Ulirsch for sharing the evaluation dataset from their publication. We also thank David Hinds, Steve Pitts, Bertram Koelsch, Michael Holmes, Stella Aslibekyan, and Nick Eriksson for their comments on the manuscript. The authors gratefully acknowledge support from AWS for GPU computing, LLM access, and credits.

The following members of the 23andMe Research Team contributed to this study: Stella Aslibekyan, Adam Auton, Elizabeth Babalola, Robert K. Bell, Jessica Bielenberg, Ninad S. Chaudhary, Zayn Cochinwala, Sayantan Das, Emily DelloRusso, Payam Dibaeinia, Sarah L. Elson, Nicholas Eriksson, Chris Eijsbouts, Teresa Filshtein, Pierre Fontanillas, Davide Foletti, Will Freyman, Zach Fuller, Julie M. Granka, Chris German, Éadaoin Harney, Alejandro Hernandez, Barry Hicks, David A. Hinds, M. Reza Jabalameli, Ethan M. Jewett, Yunxuan Jiang, Sotiris Karagounis, Lucy Kaufmann, Matt Kmiecik, Katelyn Kukar, Alan Kwong, Keng-Han Lin, Yanyu Liang, Bianca A. Llamas, Aly Khan, Steven J. Micheletti, Matthew H. McIntyre, Meghan E. Moreno, Priyanka Nandakumar, Dominique T. Nguyen, Jared O’Connell, Steve Pitts, G. David Poznik, Alexandra Reynoso, Shubham Saini, Morgan Schumacher, Leah Selcer, Anjali J. Shastri, Jingchunzi Shi, Suyash Shringarpure, Keaton Stagaman, Teague Sterling, Qiaojuan Jane Su, Joyce Y. Tung, Susana A. Tat, Vinh Tran, Xin Wang, Wei Wang, Catherine H. Weldon, Amy L. Williams, Peter Wilton. S.S., W.W., S.K., X.W., A.R., A.A., A.K., are employed by and hold stock or stock options in 23andMe, Inc.

## Data availability

Processed datasets used in our analysis, as well as the prediction results from all methods on all datasets, intermediate outputs like gene and phenotype embeddings are available via Zenodo (doi: 10.5281/zenodo.11391053).

## Code availability

Scripts used to query the LLM, as well as the scripts we use to compute our evaluation metrics are available via Zenodo (doi: 10.5281/zenodo.11391053). All prompts we used are included in the manuscript or supplementary materials.

## References

1. Kafkas, Ş., Dunham, I. & McEntyre, J. Literature evidence in open targets - a target validation platform. J. Biomed. Semant. 8, 20 (2017).

2. Tirunagari, S. et al. Lit-OTAR Framework for Extracting Biological Evidences from Literature. Preprint at 10.1101/2024.03.06.583722 (2024).

3. Sarwal, V. et al. BioLLMBench: A Comprehensive Benchmarking of Large Language Models in Bioinformatics. Preprint at 10.1101/2023.12.19.572483 (2023).

4. Chen, Y. & Zou, J. GenePT: A Simple But Effective Foundation Model for Genes and Cells Built From ChatGPT. Preprint at 10.1101/2023.10.16.562533 (2024).

5. Singhal, K. et al. Large language models encode clinical knowledge. Nature 620, 172–180 (2023).

6. Hou, W. & Ji, Z. Assessing GPT-4 for cell type annotation in single-cell RNA-seq analysis. Nat. Methods 1–4 (2024) doi:10.1038/s41592-024-02235-4.

7. Tu, T. et al. Genetic Discovery Enabled by A Large Language Model. Preprint at 10.1101/2023.11.09.566468 (2023).

8. OpenAI et al. GPT-4 Technical Report. Preprint at 10.48550/arXiv.2303.08774 (2024).

9. Anthropic. The Claude 3 Model Family: Opus, Sonnet, Haiku. Preprint at https://www-cdn.anthropic.com/de8ba9b01c9ab7cbabf5c33b80b7bbc618857627/Model_Card_Claude_3.pdf (2024).

10. Weeks, E. M. et al. Leveraging polygenic enrichments of gene features to predict genes underlying complex traits and diseases. Nat. Genet. 55, 1267–1276 (2023).

11. Stacey, D. et al. ProGeM: a framework for the prioritization of candidate causal genes at molecular quantitative trait loci. Nucleic Acids Res. 47, e3–e3 (2019).

12. Brown, T. B., et al. Language Models are Few-Shot Learners. Preprint at 10.48550/arXiv.2005.14165 (2020).

13. OpenAI et al. GPT-4o System Card. Preprint at 10.48550/arXiv.2410.21276 (2024).

14. OpenAI et al. OpenAI o1 System Card. Preprint at 10.48550/arXiv.2412.16720 (2024).

15. Team, G., et al. Gemini 1.5: Unlocking multimodal understanding across millions of tokens of context. Preprint at 10.48550/arXiv.2403.05530 (2024).

16. Grattafiori, A., et al. The Llama 3 Herd of Models. Preprint at 10.48550/arXiv.2407.21783 (2024).

17. Sclar, M., Choi, Y., Tsvetkov, Y. & Suhr, A. Quantifying Language Models’ Sensitivity to Spurious Features in Prompt Design or: How I learned to start worrying about prompt formatting. Preprint at 10.48550/arXiv.2310.11324 (2024).

18. OpenAI. New embedding models and API updates. New embedding models and API updates https://openai.com/index/new-embedding-models-and-api-updates/ (2024).

19. Mikolov, T., Chen, K., Corrado, G. & Dean, J. Efficient Estimation of Word Representations in Vector Space. Preprint at http://arxiv.org/abs/1301.3781 (2013).

20. Mountjoy, E. et al. An open approach to systematically prioritize causal variants and genes at all published human GWAS trait-associated loci. Nat. Genet. 53, 1527–1533 (2021).

21. Bordt, S., Nori, H., Rodrigues, V., Nushi, B. & Caruana, R. Elephants Never Forget: Memorization and Learning of Tabular Data in Large Language Models. Preprint at 10.48550/arXiv.2404.06209 (2024).

22. Chase, H. LangChain. (2022).

23. Khattab, O., et al. DSPy: Compiling Declarative Language Model Calls into Self-Improving Pipelines. Preprint at 10.48550/arXiv.2310.03714 (2023).

24. Pedregosa, F. et al. Scikit-learn: Machine Learning in Python. J. Mach. Learn. Res. 12, 2825–2830 (2011).

