## Supplementary Materials for "Large language models identify causal genes in complex trait GWAS"

### Supplementary Notes

#### OpenTargets and Pharmaprojects datasets

The OpenTargets gold-standard dataset is based on GWAS loci for which there is high confidence (through various sources, including metabolite GWAS, targets of existing drugs that overlap with GWAS loci, and other relevant criteria) in the causal gene. We downloaded the OpenTargets gold-standard dataset from <https://github.com/opentargets/genetics-gold-standards/> (file: <https://github.com/opentargets/genetics-gold-standards/blob/master/gold_standards/processed/gwas_gold_standards.191108.tsv>). We subsetted the dataset to only rows that had a high-confidence annotation (highest_confidence = "High"). We added gene symbol annotations to the dataset with GENCODE release 43, and excluded rows where a gene symbol was not found. This resulted in a dataset with 851 rows. To create descriptions of the phenotypes, we combined trait information from standard trait names and reported trait names. We appended reported trait names to standard trait names if the reported trait names are non-redundant, and we also removed irrelevant text from reported trait names, such as “[EA]”, “non-cancer illness code, self-reported”, “GWAS/Metabochip 2012” and “conditional on rs7709212”.

The Pharmaprojects dataset contains drug targets and indications, as well as their drug development stage. We used the Pharmaprojects dataset released by Minikel et al.^16^ and created additional mappings of disease indications to EFO (Supplementary Notes). We subsetted the dataset to only rows that have a mapped MeSH or EFO term and that correspond to launched drugs (hcat = “Launched”). We added gene symbol annotations to the dataset with GENCODE release 43, and excluded rows where a gene symbol was not found. This resulted in a dataset with 1692 causal gene - phenotype pairs. Since the Pharmaprojects data does not contain GWAS information, we created synthetic GWAS hits near the target gene for each row to mimic real GWAS hits in terms of the distance between the GWAS hit and the underlying causal gene. More details about the methods for creation of the synthetic GWAS hits can be found in the Supplementary Note.

#### OpenTargets text-mining data

The OpenTargets text mining data was obtained from the evidence table in the v23.12 OpenTargets Platform database. The evidence table contains various forms of evidence of an association between a gene and a phenotype. Specifically, the Europe PMC evidence type was retrieved from this table, which pertains to a scoring mechanism for co-occurrences of gene names and phenotypes with named entity recognition in Europe PubMed Central.

#### Pharmaprojects data processing

MeSH mappings provided by Citeline for Pharmaprojects indications were directly used to map to EFO terms using the OxO database^1^. MeSH mappings^2^ were also used to cross reference to EFO terms, followed by additional manual curation.

#### Pharmaprojects synthetic GWAS hits

The Pharmaprojects data contains only (target gene, indication) pairs and does not directly include GWAS hits. To test methods which relied on the presence of GWAS hits, we created synthetic GWAS hits near the target gene by generating lead variants. To match the features of real GWAS hits, we generated lead variant positions such that the rank distribution of causal genes from the lead variant matched that seen in real data.

We used the following procedure to generate synthetic lead variant positions:

1. We first identified the midpoint of the causal gene (target gene from Pharmaprojects).
2. We obtained a list of genes within 500 kbp of the target gene midpoint, sorted in increasing order of distance of the gene body from the midpoint of the causal gene. This list was truncated to the 20 closest genes if it had more than 20 genes.
3. We created a multinomial distribution that assigned an initial probability of 0.5 to the closest gene, 0.15 to the 2nd closest gene, 0.1 to the 3rd closest gene, 0.03 each to the 4th, 5th and 6th closest genes, and the remaining 0.16 split equally over the remaining 14 genes. If the number of genes was less than 20, the distribution was truncated to only the available genes and re-normalized to sum to 1. We chose the initial values of the probability density to approximately match the enrichment distribution of rare variant associations in genes near GWAS loci^3^ and the rank distribution of causal genes by distance described by Weeks et al.^4^
4. We selected the gene to contain the synthetic hit lead variant by sampling from the multinomial distribution.
5. We chose the midpoint of the sampled gene as the location of the synthetic hit lead variant.

This creates GWAS hits that approximately match the distribution of the rank of the causal gene in terms of gene body distance from the hit seen in real data. However, because we choose the index variant to lie within gene bodies, this is likely to create biased distributions of other genomic features corresponding to the synthetic hits.

Since the Pharmaprojects data has no underlying GWAS hits, we only evaluated “nearest gene”, text mining, and the LLM-based approaches on this dataset. For these methods, the only relevant genomic feature is the distance between the lead variant and the causal genes, therefore our results are unlikely to be strongly affected by this bias.

#### Contamination checks

One concern when evaluating performance of LLMs on novel tasks such as identifying the causal gene from a set is the effect of dataset memorization on generalization. For example, if an evaluation dataset was already seen by an LLM during training, it might perform well at identifying causal genes in that dataset, but fail to identify causal genes in a new dataset. Such contamination of the LLM training dataset by evaluation datasets is undesirable and detecting contamination can help use appropriate evaluation datasets for reporting performance on a task.

Previous attempts to detect contamination have mostly focused on detecting presence of text documents in training datasets. Recently, a method has been proposed to detect whether a tabular dataset has been seen by LLMs during training ^5^, which is more suitable for our setting. We used the tabmemcheck python package developed by the authors of Bordt et al. to evaluate if we could detect any contamination for any of our evaluation datasets.

For detecting contamination, we used the row completion tests from the tabmemcheck package on all our evaluation datasets, evaluating whether the LLM could exactly predict random rows from our evaluation datasets. Since the package requires the original forms of the dataset formatted as a CSV file, we used original versions of the dataset instead of our filtered and processed versions. As a positive control example, i.e,, a dataset which is known to be in the LLM training data, we included the UCI Iris dataset (<https://archive.ics.uci.edu/dataset/53/iris>), which is widely used in machine learning and is available on the internet. For testing, we set the few-shot parameter to 5, except for OpenTargets and GWAS catalog, where we set it to 2 due to the large number of columns. We used 25 queries to estimate the proportion of correctly predicted rows.

Figures

Supplementary Figure 1: Study design schematic showing the evaluation datasets and the methods tested on each dataset.

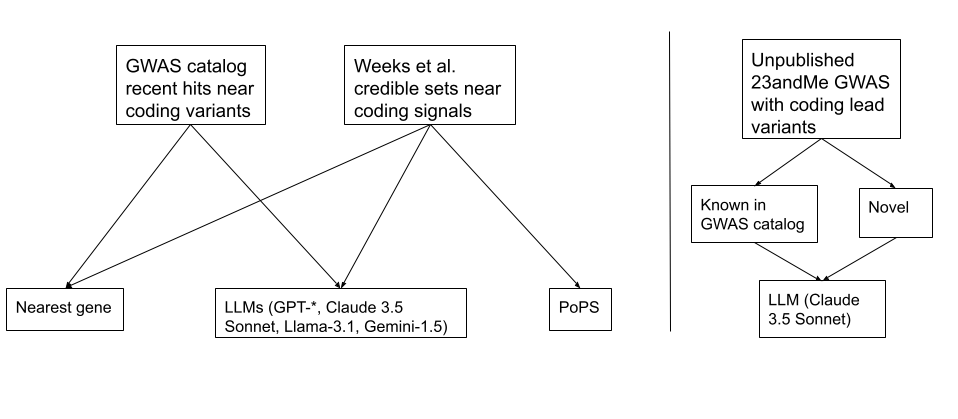

Supplementary Figure 2: Performance of all LLMs and non-LLM methods on the evaluation datasets in terms of F-score.

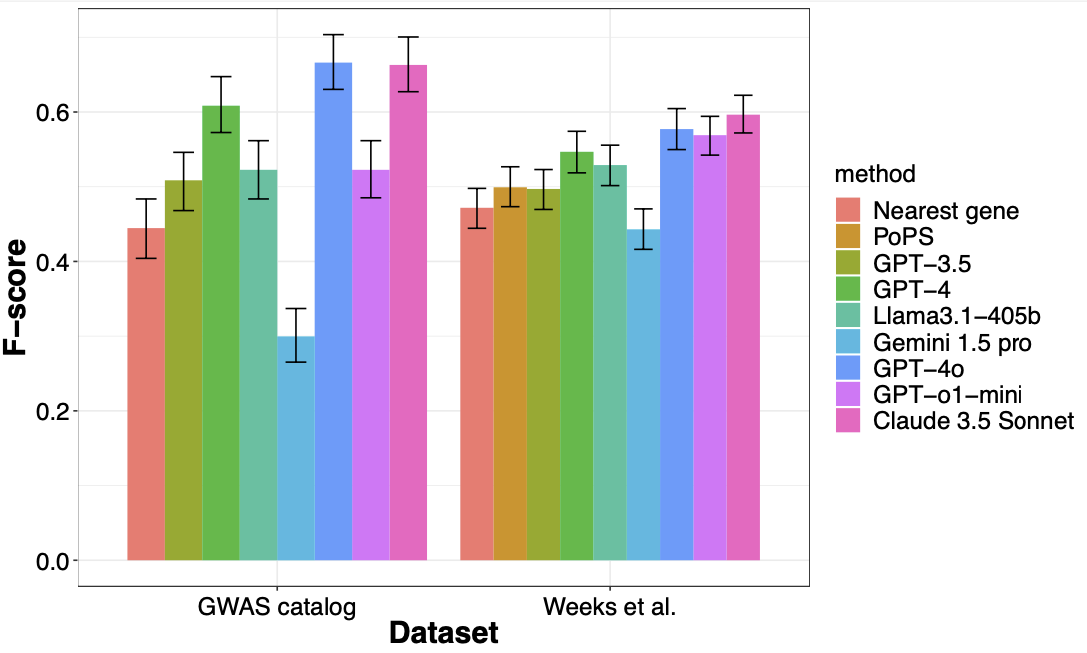

Supplementary Figure 3. Performance of methods stratified by number of genes in the locus. The left facet indicates the performance on the half where the number of genes at a locus is less than or equal to the median number for the corresponding dataset, and the right facet shows performance on the remaining half. We find that the performance gap between LLMs and other methods is magnified in gene-dense loci.

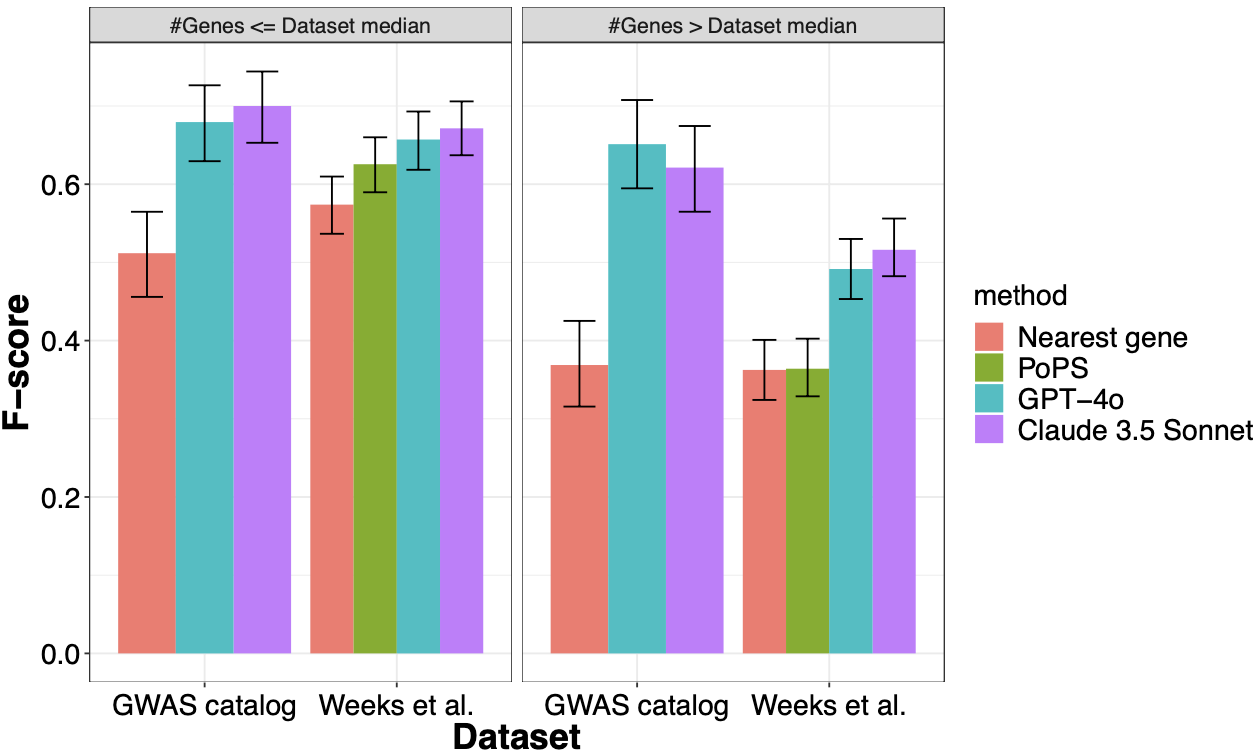

Supplementary Figure 4: Performance of two best-performing LLMs and other methods on deduplicated datasets.

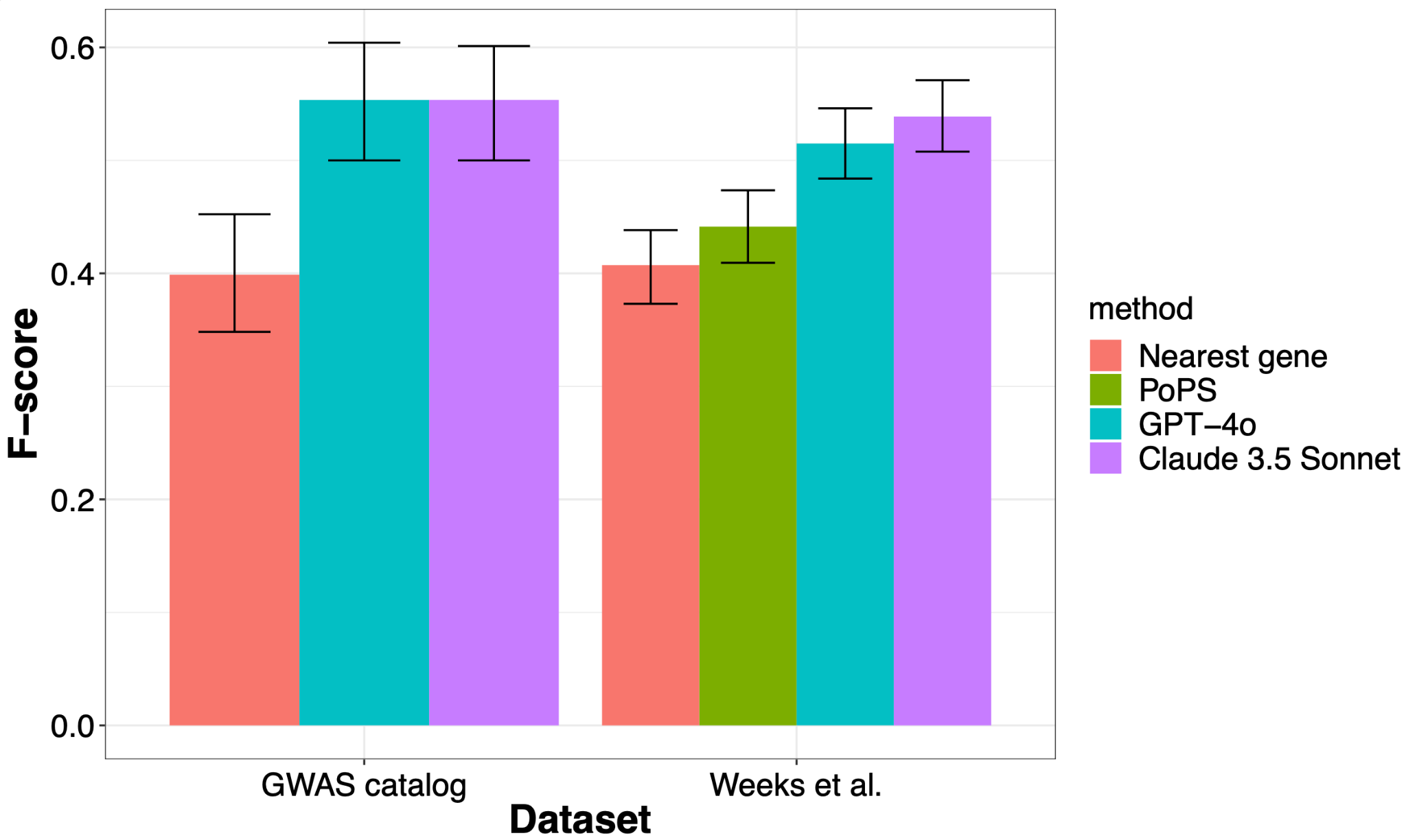

Supplementary Figure 5: Calibration plots for LLM-based approaches to assess calibration between predicted confidence and precision on all datasets

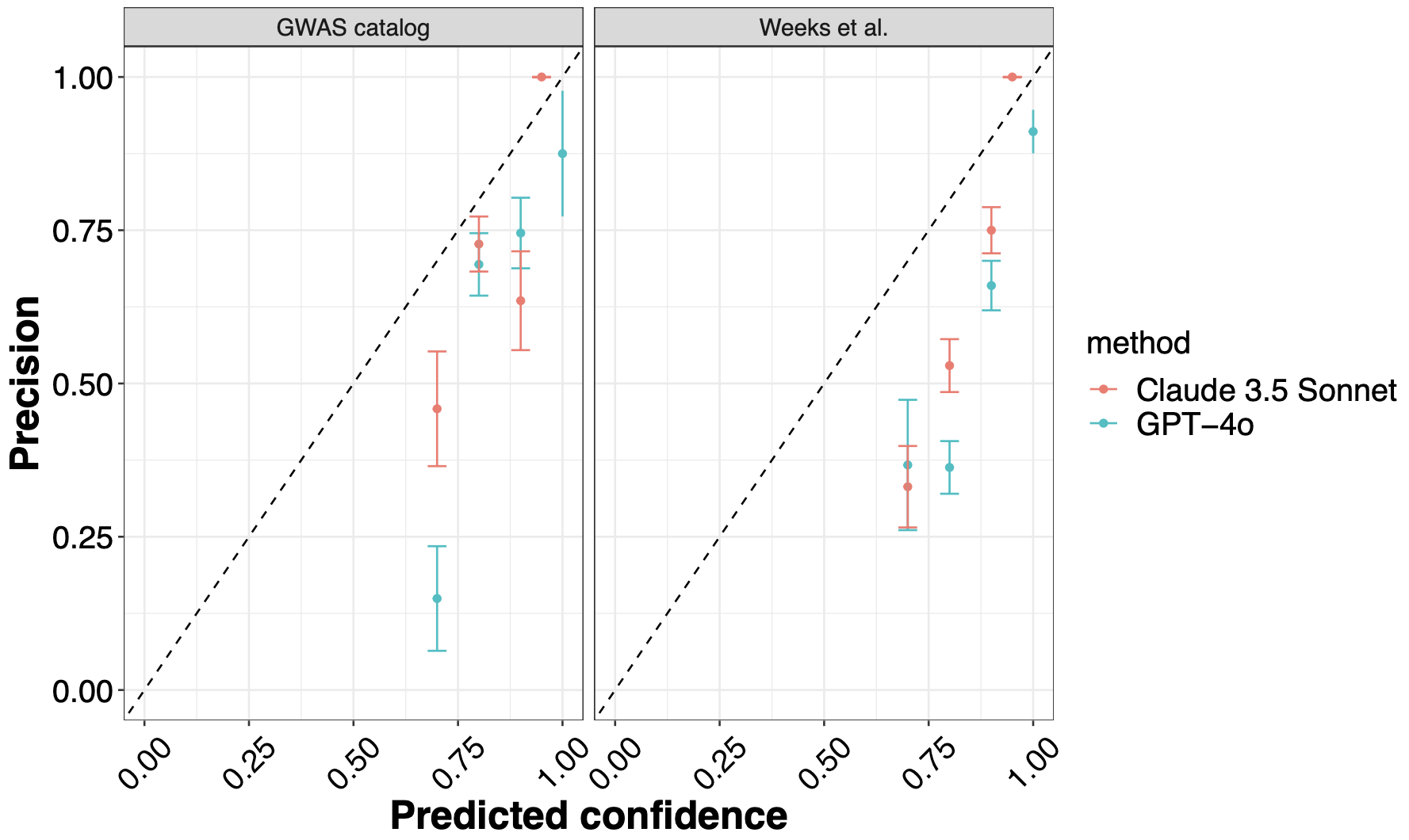

Supplementary Figure 6: Sensitivity analysis to evaluate impact of prompt format on LLM performance for Claude-3.5 Sonnet.

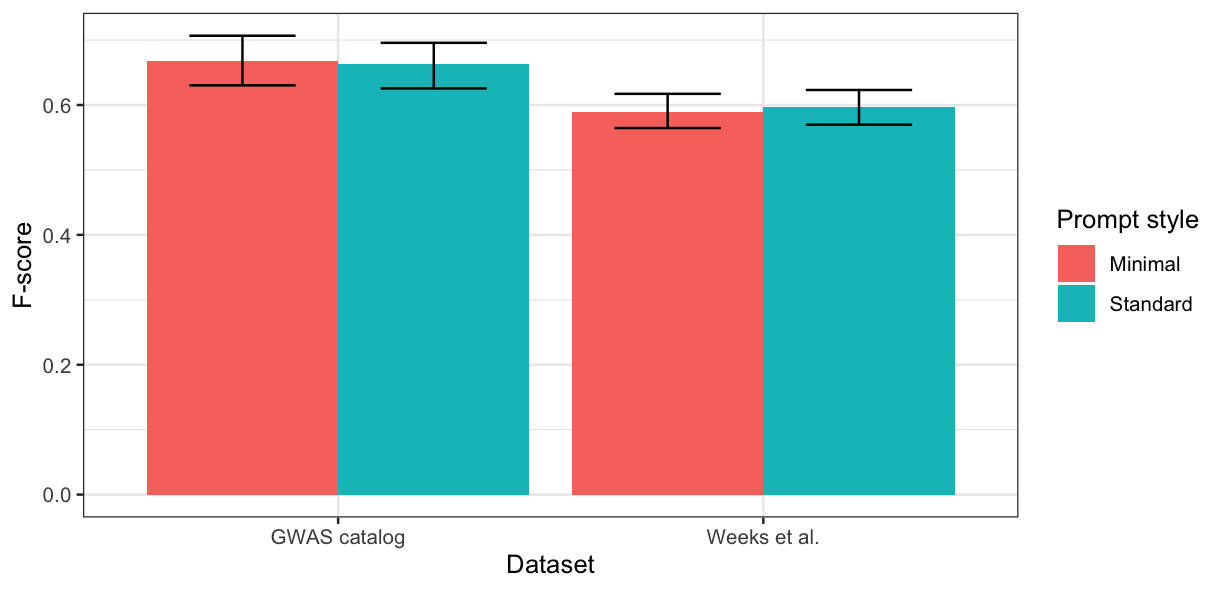

Supplementary Figure 7: Performance of embedding-based causal gene identification compared to Claude 3.5 Sonnet

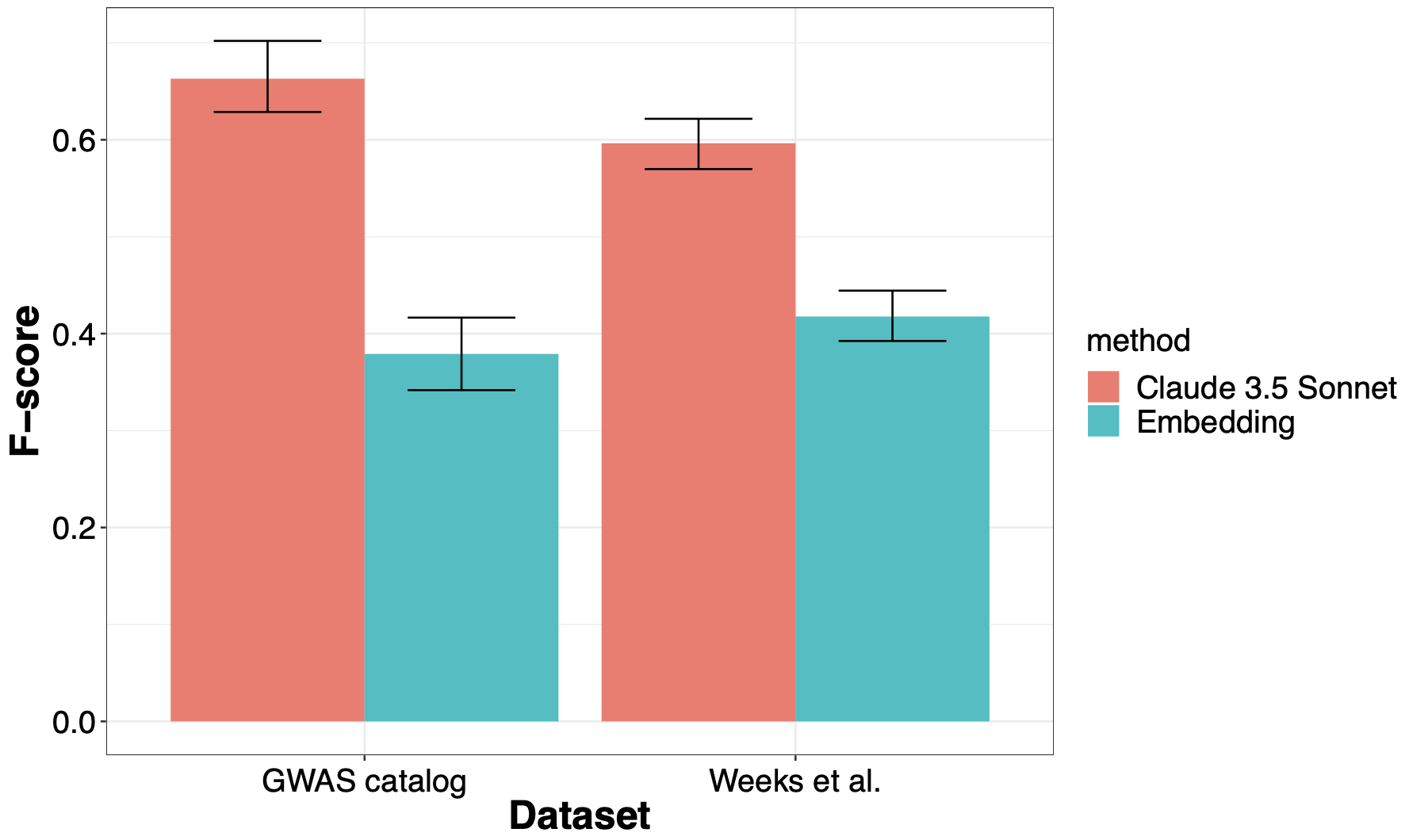

Supplementary Figure 8: Cosine similarity between the embeddings of the LDL cholesterol phenotype and genes in the PCSK9 locus from the Weeks et al. data

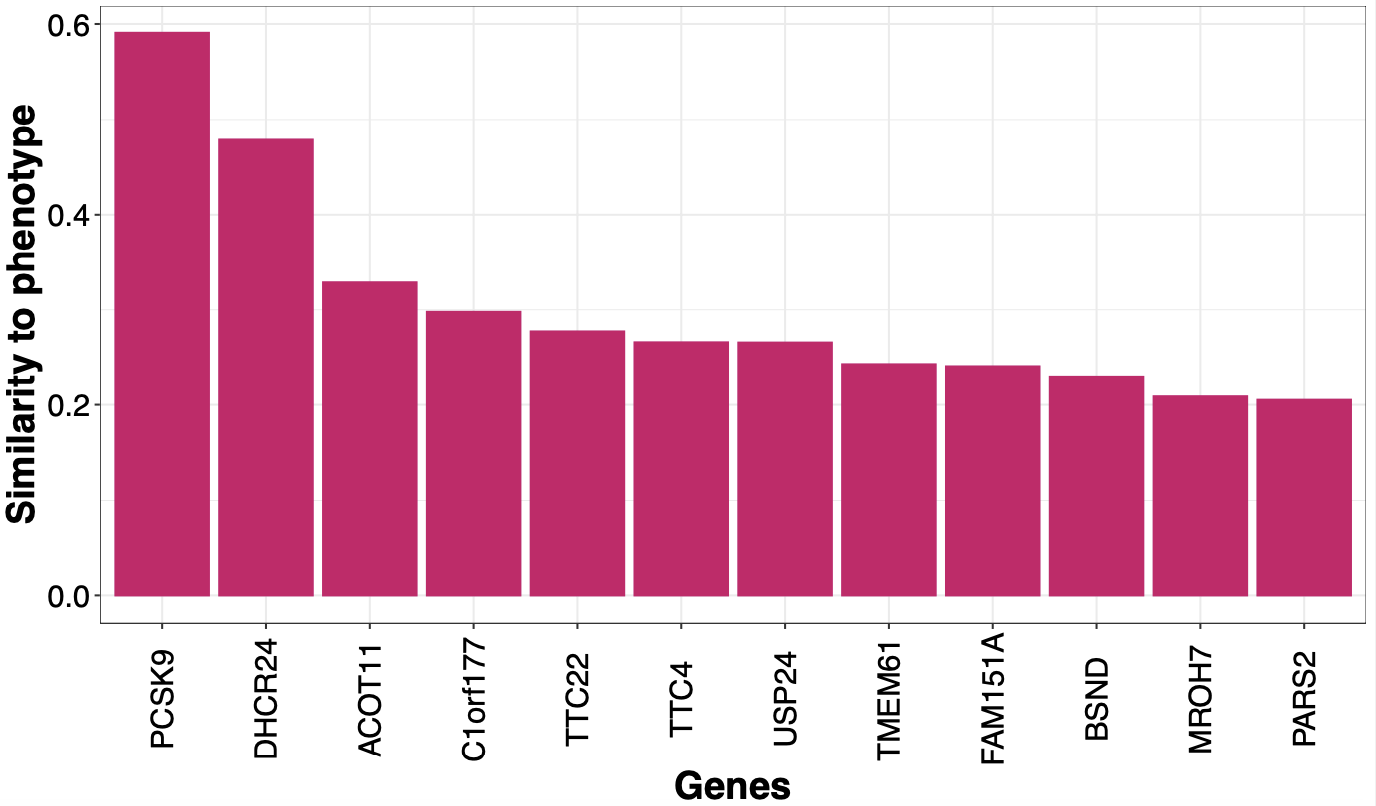

Supplementary Figure 9: Proportion of examples for each dataset where the causal gene is among the top-K (K=1 to 5) most similar genes to the phenotype in the embedding space.

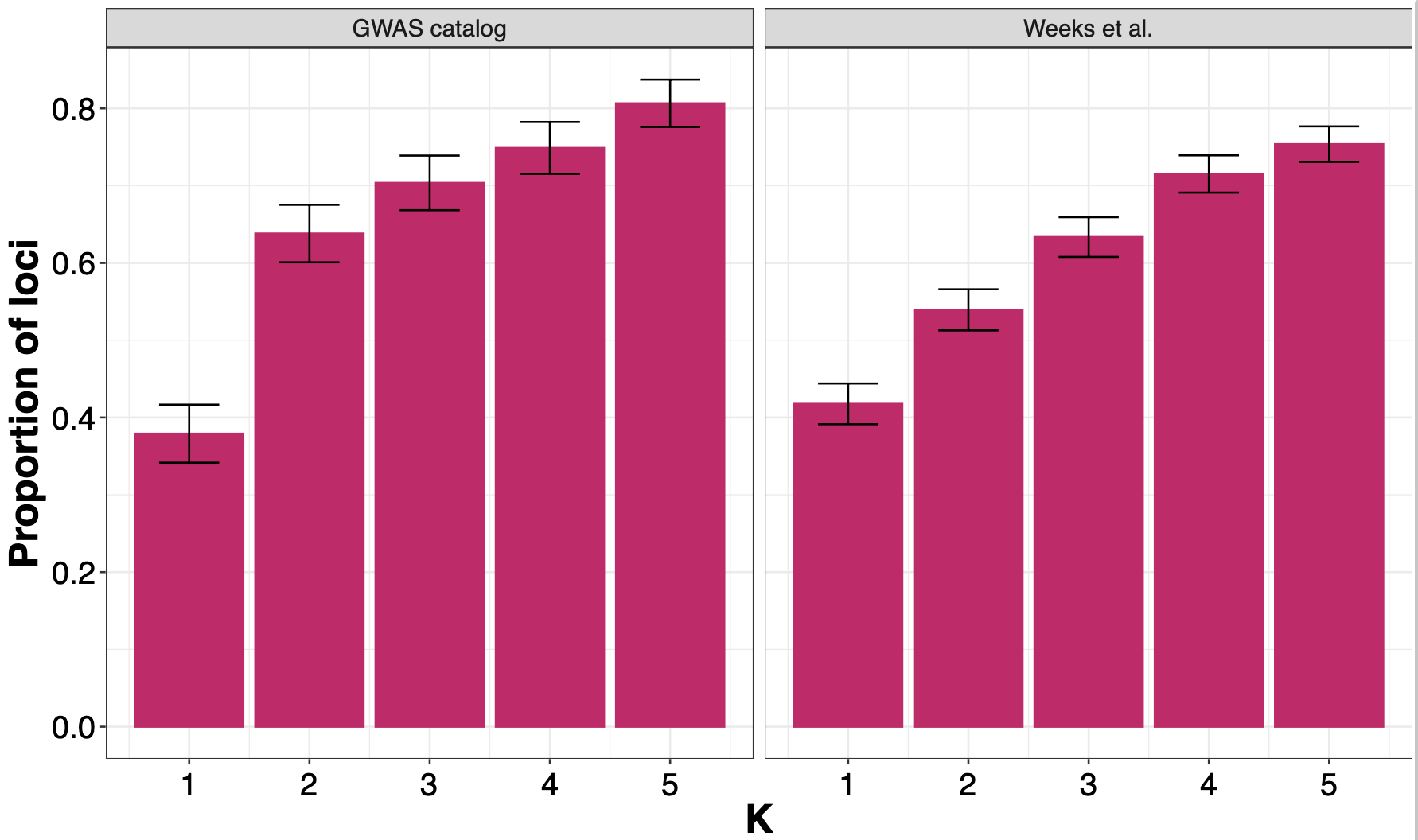

Supplementary Figure 10: Precision of LLM predictions on a larger evaluation set of 2500 loci from unpublished GWAS with linked coding variants. (a) The X-axis shows loci categorized by the maximum r-squared of the GWAS lead variant with any variant in the GWAS catalog, with increasingly lenient thresholds for the definition of novelty. The Y-axis shows precision, with 95% confidence intervals for the bars. The solid line represents the performance of LLMs at known loci, with the dashed lines indicating the 95% CI for that estimate. We see a small drop in performance for novel loci compared to known loci. (b) Precision plots stratified by LLM confidence, with each facet representing a unique confidence level for the LLM output. When stratified by confidence, the gap between performance for known and novel loci is smaller than in the aggregated data.

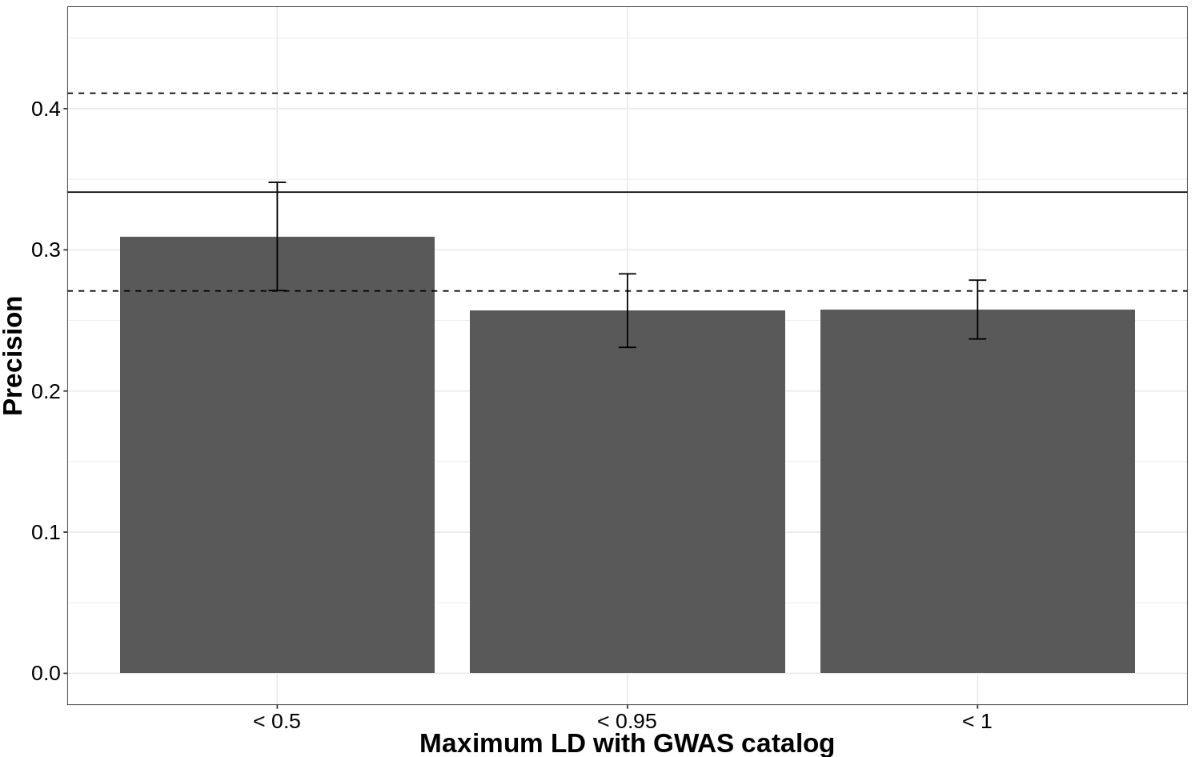

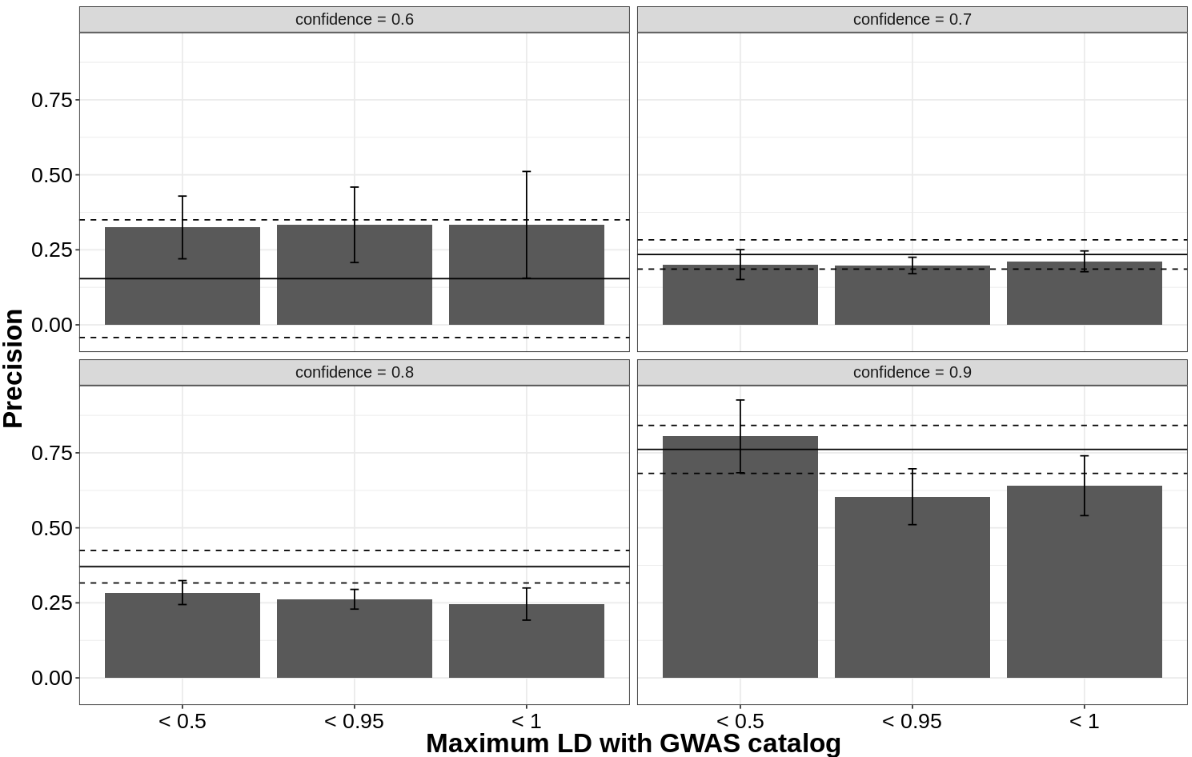

Supplementary Figure 11. Performance of chain-of-thought prompting compared to standard prompting.

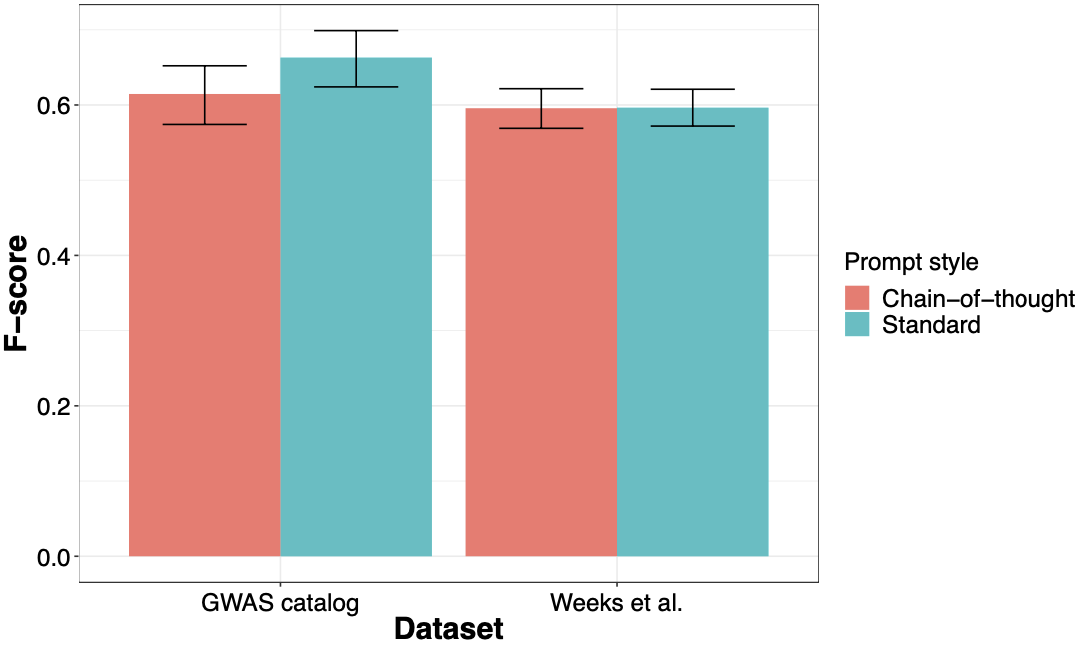

Supplementary Figure 12: Concordance between predictions from methods on all the datasets. The size indicates the proportion of loci in the dataset where both methods made a prediction, and the color indicates the proportion of agreement at loci where both methods made a prediction.

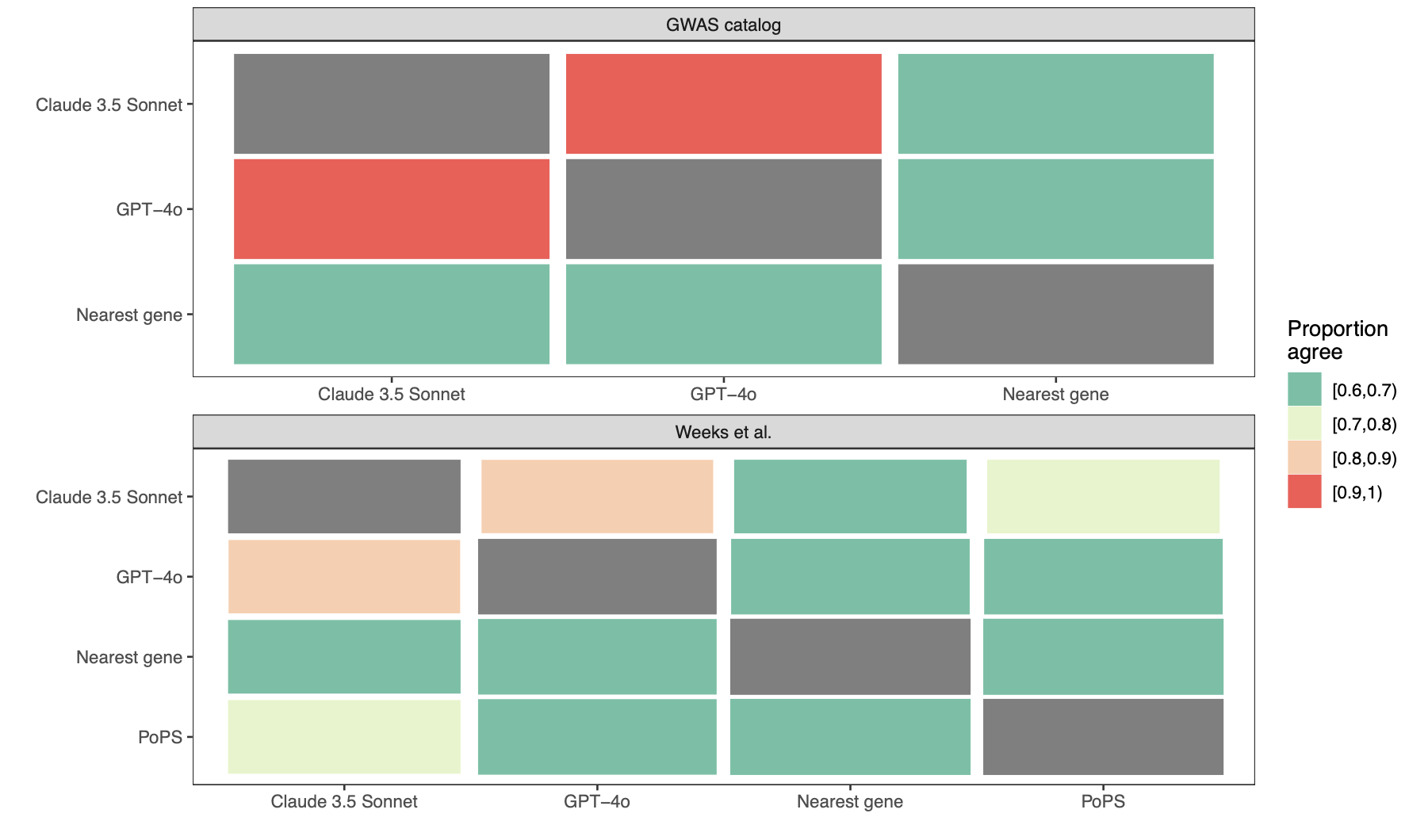

Supplementary Figure 13: Performance of the best-performing LLMs on the OpenTargets, Pharmaprojects, GWAS catalog and Weeks et al. datasets. The performance of the LLM methods is higher on the two datasets (OpenTargets and Pharmaprojects) that are easily accessible on the internet.

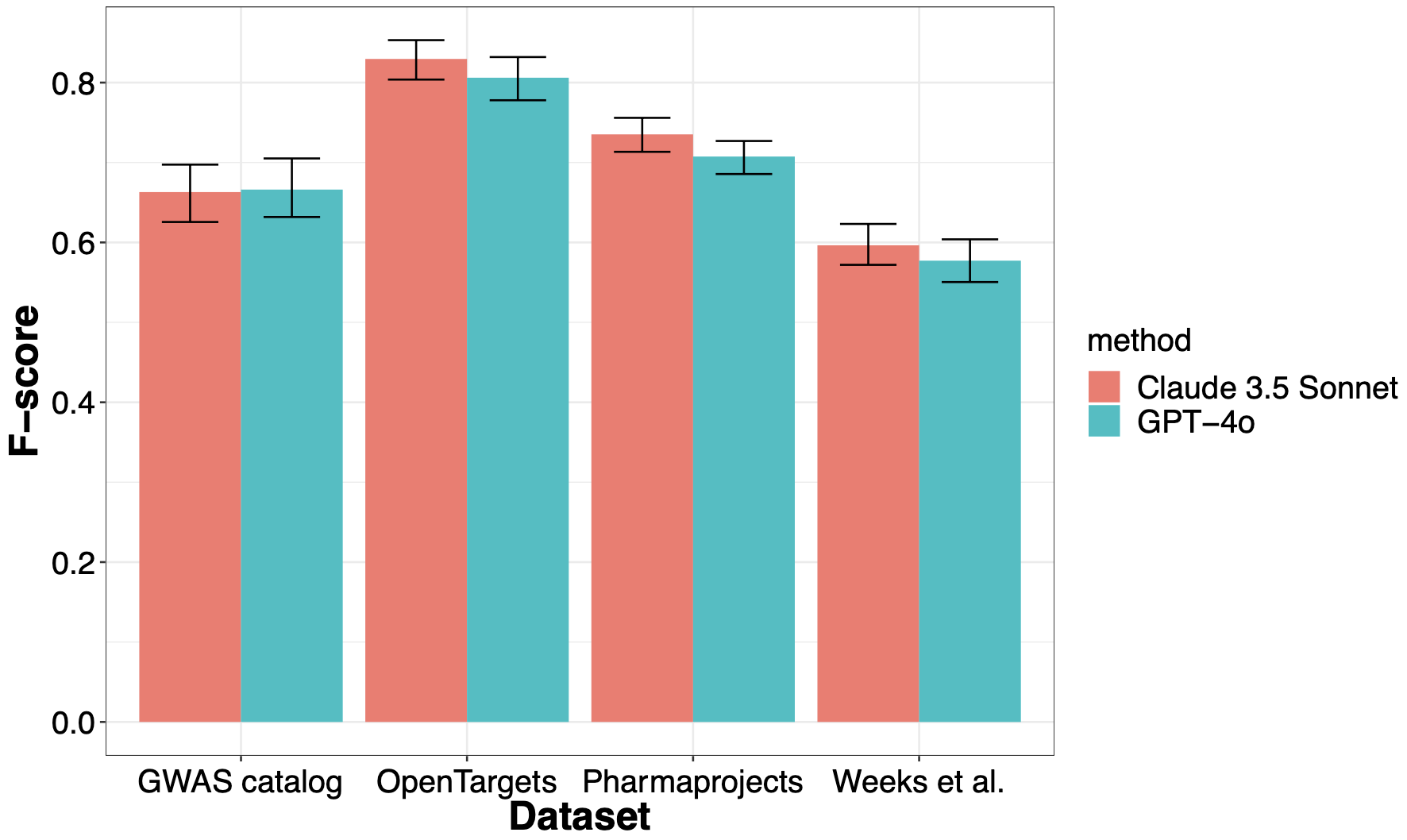

### References

1.	Jupp, S. *et al.* OxO - A Gravy of Ontology Mapping Extracts. in *Proceedings of the 8th International Conference on Biomedical Ontology (ICBO 2017), Newcastle-upon-Tyne, United Kingdom, September 13th - 15th, 2017* (eds. Horridge, M., Lord, P. & Warrender, J. D.) vol. 2137 (CEUR-WS.org, 2017).

2.	Minikel, E. V., Painter, J. L., Dong, C. C. & Nelson, M. R. Refining the impact of genetic evidence on clinical success. *Nature* (2024) doi:10.1038/s41586-024-07316-0.

3.	Backman, J. D. *et al.* Exome sequencing and analysis of 454,787 UK Biobank participants. *Nature* **599**, 628–634 (2021).

4.	Weeks, E. M. *et al.* Leveraging polygenic enrichments of gene features to predict genes underlying complex traits and diseases. *Nat. Genet.* **55**, 1267–1276 (2023).

5.	Bordt, S., Nori, H., Rodrigues, V., Nushi, B. & Caruana, R. Elephants Never Forget: Memorization and Learning of Tabular Data in Large Language Models. Preprint at https://doi.org/10.48550/arXiv.2404.06209 (2024).
